# Climate-driven rodent infection dynamics align with Lassa fever seasonality in humans

**DOI:** 10.1101/2025.10.14.25337974

**Authors:** Gregory C. Milne, Lauren A. Attfield, Joachim Mariën, Lucinda Kirkpatrick, Herwig Leirs, Kate E. Jones, Christl A. Donnelly, David W. Redding

**Affiliations:** Science Department, Natural History Museum, London SW7 5BD, UK; Environmental Change Institute, University of Oxford, 3 South Parks Road, Oxford OX1 3QY, UK; Leverhulme Centre for Nature Recovery, University of Oxford, South Parks Road, Oxford OX1 3QY, UK; Evolutionary Ecology Group, University of Antwerp, Universiteitsplein 1, 2610 Antwerp, Belgium; Virus Ecology Group, Institute of Tropical Medicine, Nationalestraat 155, 2000 Antwerp, Belgium; School of Environmental and Natural Sciences, Bangor University, Bangor, Wales; Department of Genetics, Evolution and Environment, Gower Street, University College London, London WC1E 6BT, UK; Department of Statistics, University of Oxford, 24-29 St Giles’, Oxford OX1 3LB, UK; Department of Infectious Disease Epidemiology, Imperial College London, School of Public Health, White City Campus, 90 Wood Lane, London W12 0BZ, UK

**Keywords:** Zoonotic disease, rodent-borne disease, Lassa fever, population dynamics

## Abstract

Lassa fever (LF), caused by Lassa mammarenavirus (LASV), imposes a major public health burden in West Africa. The Natal multimammate mouse (*Mastomys natalensis*, MN) is the principal reservoir, yet the extent to which host dynamics shape human outbreaks remains unclear. Leveraging long-term MN capture-mark-recapture data from Tanzania (*N*=20,249 captures, 1994–2023) and daily climate records, we quantify the demographic and climatic drivers of MN population dynamics. Using serological data on the LASV-related Morogoro arenavirus (*N*=7,850 tests, 2010–2017), we parameterize a Bayesian population model to estimate that 79.1% (95% CrI 69.3–88.9%) of infected pregnancies result in vertical transmission. Rainfall was the strongest predictor of rodent recruitment and subsequent spikes in arenavirus exposure through both vertical and horizontal transmission. When recalibrated to the Nigerian climate, our model predicted the approximate timing of LF outbreaks from 2018–2025 (*N*=6469 laboratory-confirmed cases), with predicted peaks occurring on average 0.92 months (95% CrI 0– 2.8) after observed outbreaks. These findings link climate-driven rodent demography to human LF dynamics while suggesting some generalizability between rodent-arenavirus systems. Our results provide a framework for understanding the drivers of rodent host population dynamics that, in turn, shape the temporal and spatial patterns of human zoonotic disease risk.

**Significance Statement:** Lassa fever (LF), caused by Lassa mammarenavirus, remains a major public health threat in West Africa. Since human infections arise as spillovers from the Natal multimammate mouse (*Mastomys natalensis*, MN), understanding rodent demography may clarify patterns of human risk. Using long-term data on MN populations, arenavirus exposure, and climate from Tanzania, we show that rainfall strongly predicts rodent recruitment and arenavirus exposure, with an estimated 79% of infected pregnancies resulting in vertical transmission. When recalibrated to the Nigerian climate, predicted peaks in infected rodents closely aligned with annual LF outbreaks. These results highlight how demographic and climatic factors drive rodent populations and may, in turn, shape the timing of zoonotic disease risk.

## Introduction

Rodents are a broadly distributed, highly diverse and often synanthropic order that host numerous zoonotic pathogens (1–4). Since zoonotic pathogens are maintained in and emerge from animal reservoir hosts, understanding the ecological basis of disease emergence is increasingly recognized as essential for public health and pandemic prevention (5, 6). Insights on the drivers of reservoir host population dynamics can inform disease surveillance and prevention (7), as well as both short- and long-term outbreak forecasts (8, 9).

Rodents’ “fast” life-history traits (1) allow rapid responses to fluctuations in resource availability. Their population dynamics are driven by density-dependent processes such as competition and predation, and by density-independent processes such as seasonal or interannual variation in climate that act on rodent populations either directly (e.g. through winter survival) or indirectly (e.g., via trophic cascades) (10–14). While many studies have associated past climate variability with rodent abundance or pathogen prevalence in reservoirs (8, 15–18), few have developed process-based models that link environmental drivers to host demography and infection dynamics (10). Yet such models are critical for making mechanistic, transferable predictions of zoonotic risk, beyond the range of observed data.

Developing and applying these models has broad relevance beyond rodent systems. Process-based frameworks that integrate host demography, infection, and climate can improve our understanding of transmission dynamics in a wide range of zoonotic systems—whether the hosts are rodents, bats, birds, or ungulates. By estimating key demographic and transmission parameters and quantifying how they respond to environmental variation, these models allow attribution of climate and climate-change effects in a mechanistic rather than phenomenological way. Broader use of such approaches could therefore advance comparative insights across host-pathogen systems and improve forecasts of animal-borne disease risk.

One such climate-sensitive zoonotic disease is Lassa fever (LF), endemic to West Africa and responsible for an estimated 2.1–3.4 million human infections annually (19), most of which remain undiagnosed (20). The Natal multimammate mouse (*Mastomys natalensis*, MN), is the principal reservoir of Lassa mammarenavirus (LASV), the etiological agent of LF. LASV is one of several arenaviruses found across the six MN subtypes distributed throughout Africa (21). Most human LF cases are thought to arise from direct or indirect contact with infected MN (22) suggesting that human risk is closely tied to MN ecology. Seroprevalence studies of LASV-like arenaviruses in rodents show strong seasonal patterns (23–25), as do reported LF cases in humans (26). However, given the persistence of MN in many peridomestic settings in West Africa (27), it remains unclear whether these seasonal outbreaks reflect fluctuations in MN population dynamics and infection prevalence, seasonal changes in human behavior (e.g., agricultural activities), or both.

In Eastern and Southern Africa, MN has been extensively studied as an agricultural pest and host of several non-pathogenic arenaviruses (21, 23, 24, 28). These studies have been pivotal for understanding MN ecology and arenavirus transmission beyond Lassa-endemic regions, and have informed LASV vaccine development (29, 30), predictions of LASV distribution under anthropogenic change (31), and estimates of arenavirus shedding dynamics (24). Within Lassa-endemic West Africa, however, available MN data remain sparse (32, 33), limited by biohazard and ethical constraints on trapping LASV-positive rodents. This scarcity has hindered our ability to link host dynamics with LF outbreak timing and to build predictive, mechanistic models of human risk—models that could guide preventive measures and the allocation of health resources.

In this study, we use a recently published three-decade capture-mark-recapture (CMR) dataset of MN from Morogoro, Tanzania (28), together with paired serological data on the LASV-related Morogoro arenavirus (MORV) (34) and local climate data (35, 36), to examine the demographic and environmental drivers of MN population dynamics. We develop a Bayesian integral projection model (IPM) that jointly captures MN demography and arenavirus transmission, allowing us to quantify the relative contributions of density-dependent and climatic processes, and to estimate parameters governing vertical and horizontal transmission. We then assess whether model dynamics parameterized from East Africa can generalize to West Africa by recalibrating to Nigerian climate conditions and comparing model predictions with observed LF outbreak patterns. Through this approach, we evaluate whether mechanistic models of reservoir-pathogen systems can provide transferable insights across regions and closely related viral and host species.

## Results

### Bayesian statistical models reveal drivers of rodent demographic processes

We developed Bayesian statistical models to describe changes in MN body weight, recruitment, and survival (Fig. 1) using CMR data (*N*=20,249 captures, 1994–2023) (28) and lagged, decomposed climate variables (35, 36).

**Fig. 1.**
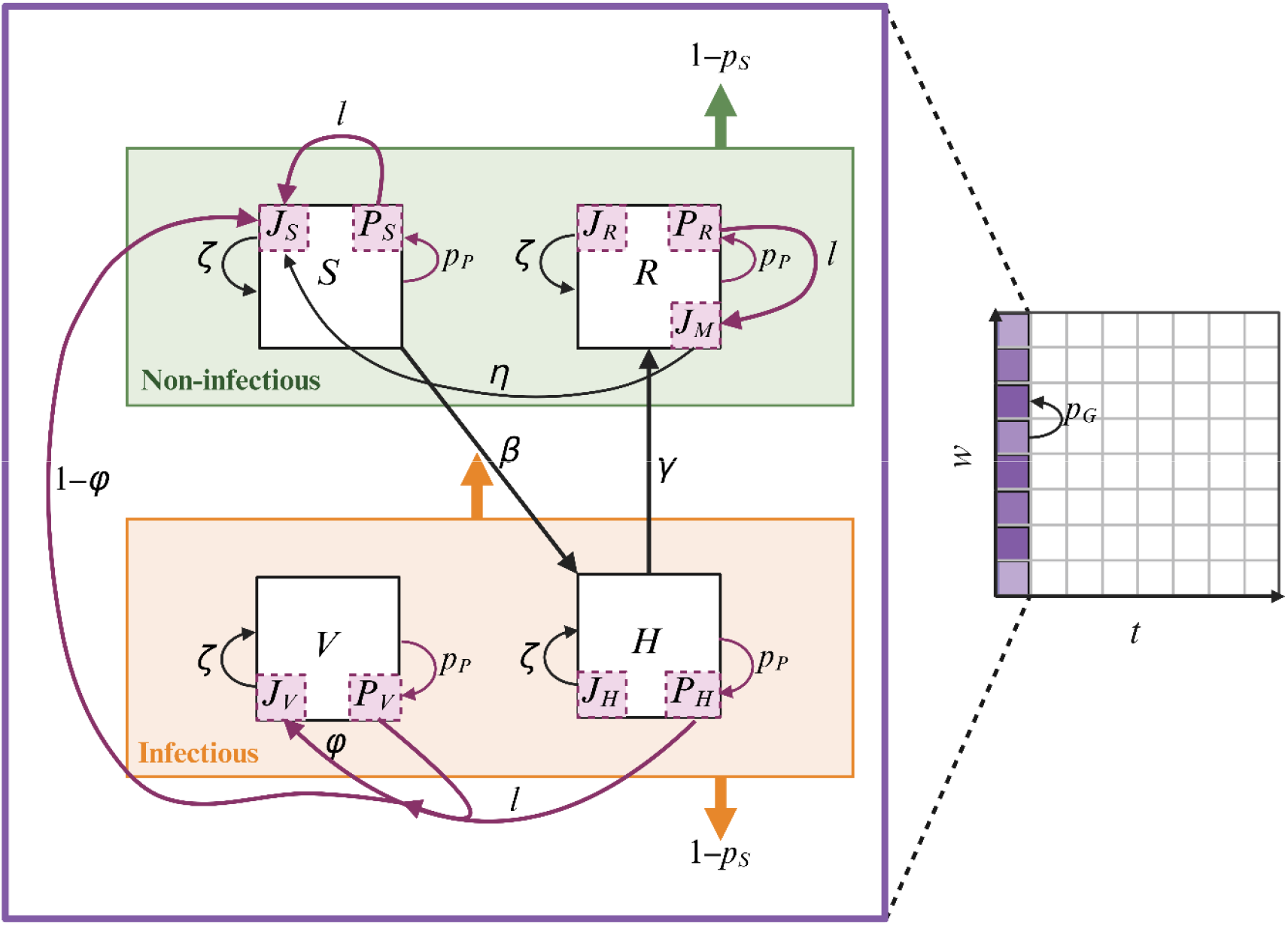
Integral projection model. The population is divided into weight classes (purple grid), age classes (juveniles and adults otherwise), and four epidemiological compartments representing susceptible (), horizontally infected (), vertically infected (), and recovered () individuals. Individuals change body weight at probability, become pregnant at probability (transitioning to sub-compartment), and die at probability. Pregnant individuals produce offspring which enter the juvenile sub-compartment (). Pregnancies from infected compartments (and) have probability of producing vertically infected juveniles (), with the remainder being susceptible (). Recovered mothers () produce juveniles in a transiently maternal antibody-positive sub-compartment (), transitioning at rate to susceptible juveniles (). Juveniles mature into adults at rate (and are thereafter able to become pregnant). Individuals in the infectious super-compartment (orange box) contribute to infection of susceptible individuals via horizontal transmission rate, with recovery from to occurring at rate.

Best-performing models included both population size and weight, along with climate predictors: body weight change—precipitation seasonality and trend; recruitment—precipitation seasonality and variability; survival—precipitation seasonality and temperature trend (Fig. 2A; *SI Appendix*, Figs. S1–S4). Models without climate predictors ranked in the lowest 12.5% (8/64) by marginal log-likelihoods (*SI Appendix*, Figs. S1–S3), underscoring the importance of climate in explaining demographic variation. No consistent climate lag structure was detected, with posterior weights relatively uniformly distributed over the 0–168-day range (*SI Appendix*, Fig. S5). Predictions from the best-performing models captured seasonal variation in all three processes (*SI Appendix*, Fig. S6–S9), indicating good model fit to observed dynamics.

**Fig. 2.**
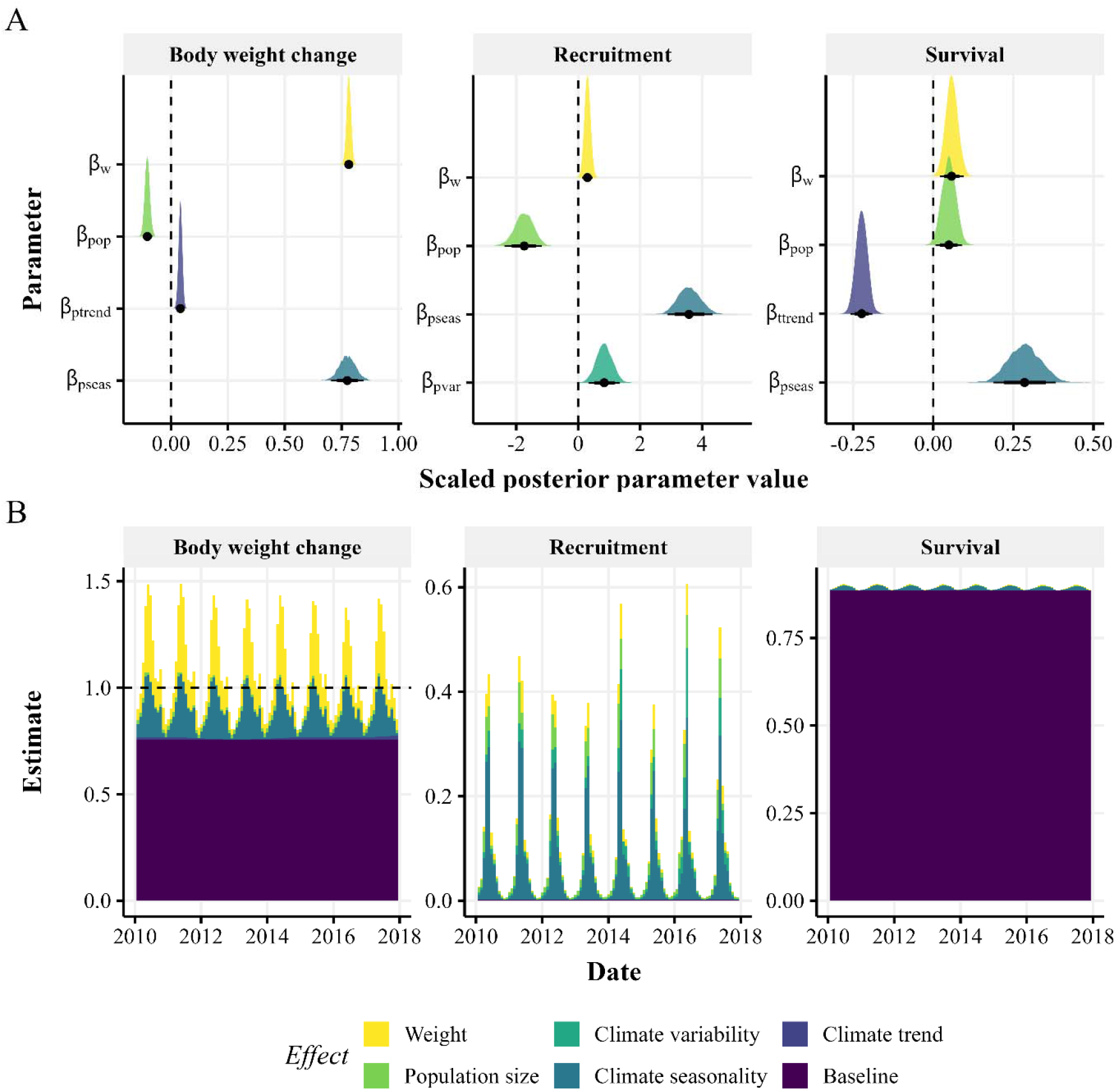
Estimating the demographic and climatic predictors of rodent demography. ***(A)*** Estimated posterior distributions of unit-scaled coefficients from demographic process models. The density of each posterior distribution is shown alongside a posterior median (point), 80% credible interval (thick horizontal line), and 95% credible interval (thin horizontal line). The vertical dashed line represents no effect. Notation: *β*_*w*_, body weight; *β*_*pop*_, population size; *β*_*ptrend*_, precipitation trend; *β*_*pseas*_, precipitation seasonality; *β*_*pvar*_, precipitation variability; *β*_*ttrend*_, temperature trend. ***(B)*** Estimates from *N*=1,000 posterior predictive simulations of the integral projection model showing the median contributions of demographic and climatic effects to each demographic process (with colors corresponding to the parameters in panel *(A)*. Bars show the time-specific contribution of each covariate to the demographic process, benchmarked against the response that would be predicted if all marginal effects took their minimum value (Baseline). For survival, this baseline value includes the proportion of rodents estimated to be alive despite never being recaptured (*SI Appendix*). The total height of stacked bars is the time-specific predicted response. Units for body weight change are the time-specific predicted weights (grams) made proportional to the mean modelled weight over time (dashed horizontal line), while units for recruitment and survival are probabilities. Demographic process models were fit to decomposed climate data (35, 36) and *M. natalensis* capture-mark-recapture data (28) using a No-U-Turn Sampler algorithm in Stan (53). Posterior predictions were generated using Bayesian melding (65) whereby demographic samples were reweighted based on their consistency with seroprevalence data (34).

Partitioning the relative contributions of predictors (Fig. 2A) showed that body weight change was primarily driven by previous weight and seasonal precipitation; recruitment by population size and seasonal precipitation (with moderate effects of precipitation variability and body weight); and survival showed limited dependence on any predictor (Table 1; Fig. 2B; *SI Appendix*, Fig. S10). These results indicate that MN population dynamics are most strongly influenced by recruitment and body weight change.

**Table 1.** The drivers of rodent demography. Percentage of the variation in modelled demographic process explained by each covariate. Estimates are given as the median posterior predictions with 95% credible intervals (*N*=1,000 posterior predictive simulations).

|  | Body weight change | Recruitment | Survival |
| --- | --- | --- | --- |
| Weight | 10.82 (1.67–27.86) | 5.86 (0.74–17.02) | 0.53 (0.11–1.05) |
| Population size | 1.34 (0.10–2.40) | 26.43 (1.77–55.11) | 0.37 (0.02–1.63) |
| Climate trend | 0.83 (0.01–1.68) | NA | 0.62 (0.04–2.99) |
| Climate seasonality | 11.42 (0.40–20.59) | 52.12 (2.11–78.96) | 4.55 (0.20–5.34) |
| Climate variability | NA | 11.46 (1.97–28.82) | NA |

604 Abbreviations: NA, not applicable (covariate not in model).

### Linking demography to infection reveals key processes driving arenavirus transmission

Fitting the demographic–infection model (Fig. 1) to MORV seroprevalence data (*N*=7,850 tests, 2010–2017) (34), yielded an estimated 79.1% (95% credible interval (CrI) 69.3–88.9%) probability of vertical transmission in infected pregnancies (Fig. 3A), indicating that this route likely plays a major role in maintaining viral persistence between breeding seasons. Model predictions reproduced observed seasonal seroprevalence patterns (Fig. 3B) and moderately explained 2017 hold-out data (posterior coefficient = 0.98, 95% CrI 0.62–1.34), outperforming an intercept-only null model (Supplementary Results).

**Fig. 3.**
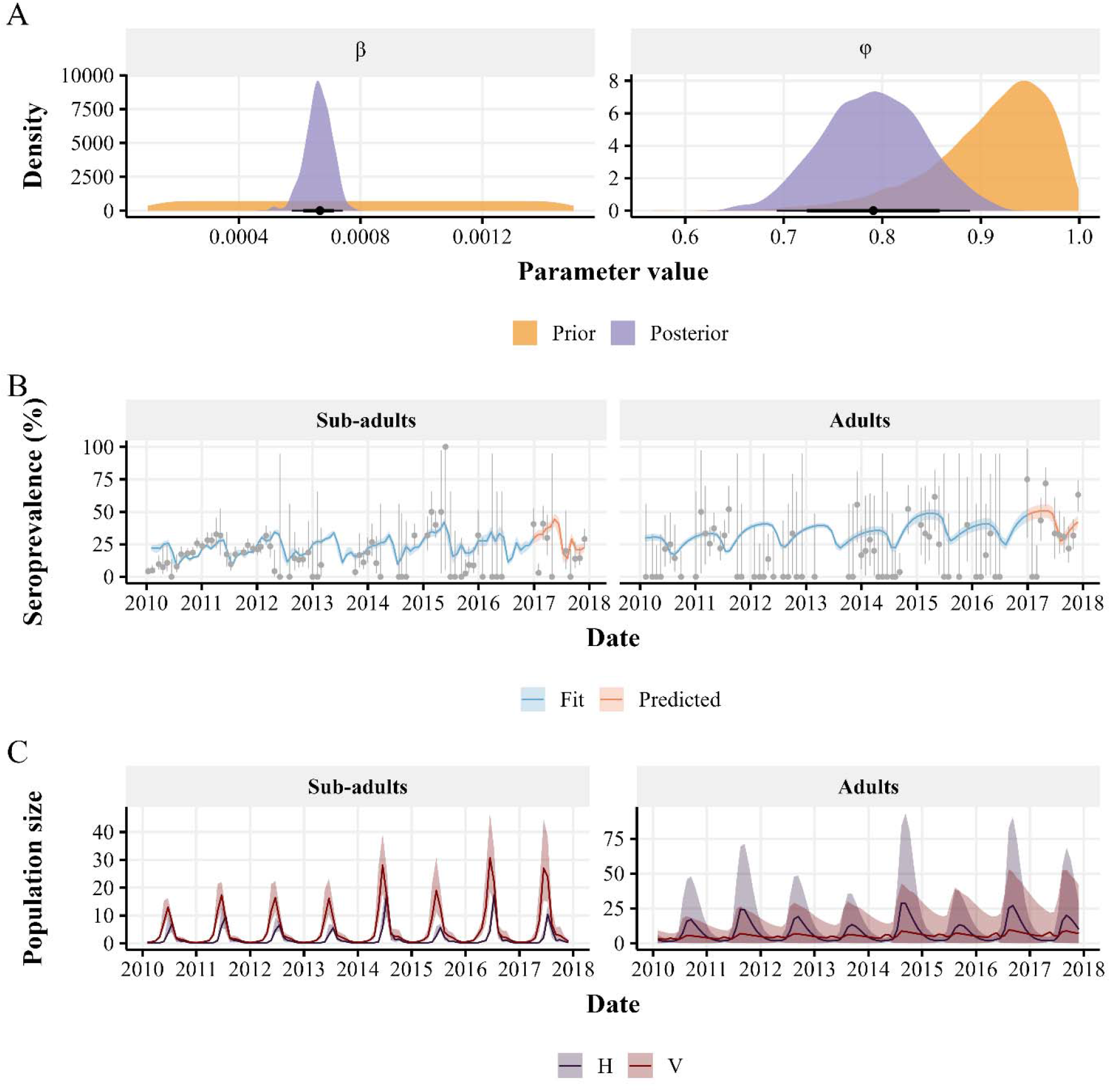
Estimating the drivers of arenavirus transmission. ***(A)*** Approximate posterior distributions of the horizontal transmission rate (*β*) and the probability of vertical transmission from an infected pregnancy (*φ*), with associated prior distributions (uniform and beta, respectively). Points represent the posterior medians, thick inner lines the 80% credible intervals, and thin outer lines the 95% credible intervals. ***(B)*** Fit of modelled seroprevalence to observed *Mastomys natalensis* Morogoro arenavirus seroprevalence data for sub-adults (*N*=6,904 tests) and adults (*N*=946 tests) (34). Points and error bars represent observed means and binomial 95% confidence intervals for each trapping session, while lines and shaded areas show posterior predictive medians and 95% credible intervals (*N*=1,000 posterior draws). The integral projection model was fit to seroprevalence data from 2010–2016 (blue), with 2017 withheld for validation (red). Posterior predictions were generated using Bayesian melding (65) whereby demographic samples were reweighted based on their consistency with seroprevalence data (34). ***(C)*** Posterior predictions (medians and 95% credible intervals) from the fitted model of sub-adult and adult population size by infection compartment: horizontally infected (*H*) and vertically infected (*V*) (*N*=1,000 posterior draws).

To identify outbreak drivers, we examined climatic and demographic conditions preceding major seroprevalence peaks in 2015 and 2017 (Fig. 3B). Unusually high rainfall during 2014 and 2016 increased recruitment probability (Fig. 2B; SI Appendix, Fig. S11), explaining, respectively, 17.1% and 17.5% of recruitment variance—substantially more than in 2013 (8.3%), 2015 (11.2%), or the overall model (11.5%; Table 1; *SI Appendix*, Fig. S10). Elevated recruitment increased vertical transmission events and, subsequently, horizontal transmission, with the 2016 peak in horizontally infected adults roughly twice that of 2015 (Fig. 3C). The resulting accumulation of maternally and postnatally acquired antibodies caused seroprevalence to peak about 6.5 months after infection prevalence (SI Appendix, Figs. S12–S13). In contrast, we found no shifts in predictors of body weight change or survival prior to these outbreaks (SI Appendix, Fig. S10). Together, these results indicate that recruitment—amplified by excess rainfall—is the dominant process driving arenavirus outbreaks.

### Rodent infection dynamics predict the timing, but not magnitude, of Lassa fever outbreaks

To test the model’s generalizability, we recalibrated it with climate data (36, 37) from the five Nigerian states with the highest LF burden (Fig. 4A) and compared predicted MN infection dynamics to reported LF cases from 2018–2025 (*N*=6,469) (26).

**Fig. 4.**
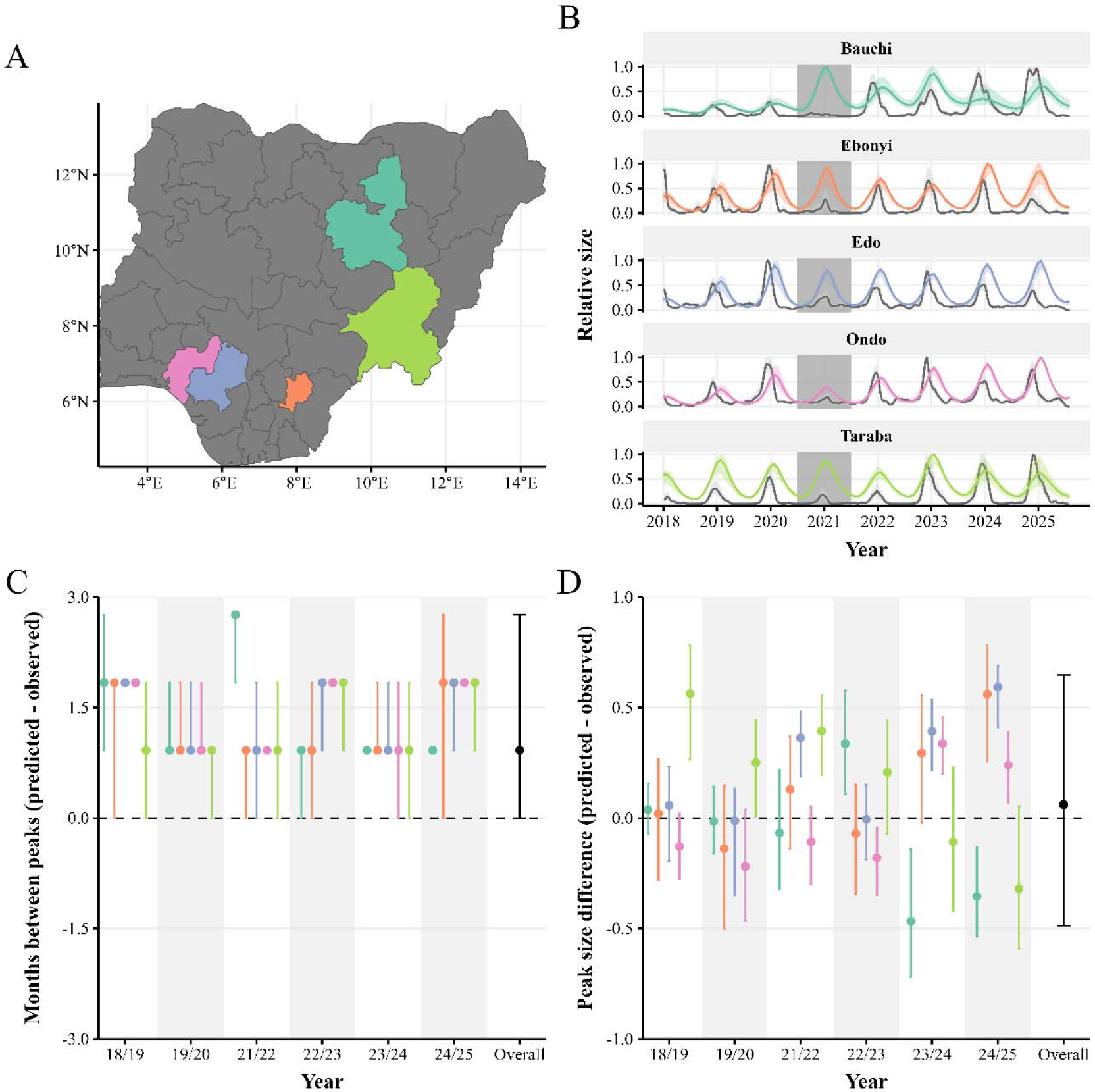
Assessing generalizability across arenavirus systems. Nigeria posterior predictive simulations (*N*=1,000) using the recalibrated model. ***(A)*** The included Nigerian states, with colors corresponding to state names in panel B. ***(B)*** Median (line) and 95% credible interval (shaded area) of the predicted relative number of infected rodents (color) versus the relative number of laboratory-confirmed Lassa fever cases transformed to infections (grey; *N*=6,469 cases) (26). Uncertainty in reported cases was represented by modelling counts with *N*=1000 random draws from a Poisson distribution parameterized with rate parameter set to the number of reported cases. Estimated dates of infection were obtained by transforming dates of case reporting by an inferred Lassa fever incubation period distribution (sampling *N*=1,000 random parameter sets; see *SI Appendix*). Predictions and inferred infections were scaled to the maximum value in each simulation. Data from the 2020/21 season were omitted from analyses due to interruptions in case reporting during the COVID-19 pandemic (26) (dark grey shaded area). ***(C–D)*** Difference in the timing ***(C)*** and size ***(D)*** of predicted and observed outbreak peaks (showing medians as points and 95% credible intervals as error bars). The dashed horizontal lines indicate perfect agreement. ‘Overall’ estimates are calculated across all years and states. Lassa fever seasons were defined as July–June to capture the typical December–April outbreak period (26) (e.g., for year 18/19 we estimated the outbreak peak occurring between July 2018 and June 2019).

Predicted infection dynamics matched the periodicity of LF outbreaks (Fig. 4B), with mean predicted peaks lagging observed peaks by 0.92 months (95% CrI 0–2.76) (Fig. 4C). Across states and years, 37% (11/30) of difference intervals overlapped zero, indicating in these instances more exact concordance between predicted and observed peak timing. This relationship is nontrivial, as the primary climatic drivers of MN demography peak at different times of year from LF outbreaks; other demographic metrics such as predicted body condition or recruitment probability were weaker predictors of outbreak timing (SI Appendix, Fig. S14).

Predicted number of rodent infections, however, did not explain LF outbreak magnitude. Across states and years, the relative size of predicted peaks exceeded observed peaks by 0.06 on average (95% CrI −0.49, 0.65) (Fig. 4D). Although performance was moderate in certain settings (e.g., Ondo state in most years), predictions of outbreak size were generally poor. These findings suggest that while modelled MN transmission dynamics capture the timing of LF outbreaks, additional ecological or behavioral factors likely determine their magnitude.

## Discussion

By combining extensive empirical data with a novel population dynamic model, we show how climate and demography shape the population and infection dynamics of a major zoonotic reservoir.

Our models identify precipitation as a key driver across MN demographic processes. Rainfall has long been linked to rodent abundance, particularly where it is a limiting factor for primary productivity and food resources (37). The positive relationship we found between recruitment and unseasonably heavy rainfall likely reflects increased resource availability, as MN breeding responds rapidly to food abundance (27, 38). Recruitment was the most climate-sensitive process, with two-thirds of its variation explained by precipitation. Given the vulnerability of Tanzania—and especially semi-arid Morogoro—to climate change (39), MN breeding and dispersal patterns are likely to become more unpredictable as shifting rainfall patterns may alter the timing or intensity of breeding and flooding could force rodent dispersal from burrows. The strong El Niño event of 2015–2016, which brought exceptional rainfall and flooding to Tanzania (40), coincided with the predicted spike in recruitment in 2016 (Fig. 2B), illustrating this link.

Body weight and population size also influenced MN demography. Heavier individuals were more likely to survive and reproduce (Fig. 2A), consistent with theory and previous studies showing that better body condition buffers against starvation and enhances fecundity (10, 13, 41). Population size had negative effects on weight gain and recruitment—likely reflecting competition and density-dependent breeding (13, 42)—but a positive effect on survival, possibly due to reduced per-capita predation risk in larger populations (43).

Our results highlight the importance of vertical transmission for arenavirus persistence. We estimate that 79.1% (95% CrI 69.3–88.9%) of infected pregnancies transmit infection to offspring (Fig. 3A), providing the first field-based estimate of this probability. Since the infectious period after acute infection is short (24), vertical transmission likely underpins long-term viral maintenance in MN populations (24, 25, 34, 44). This has significant implications for control strategies, suggesting that fertility management may effectively reduce arenavirus persistence, whereas untargeted culling is unlikely to be effective (34, 42). Our estimates align with laboratory studies of LASV (99.3%) and MORV (100%) (24, 44), with minor differences plausibly due to variation in host genetics and immune responses between wild and laboratory rodents (45).

Climatic factors also shape arenavirus outbreak dynamics. Excess rainfall increased recruitment, which elevated vertical transmission and, through the influx of naïve juveniles, subsequent horizontal transmission (Fig. 3B; SI Appendix, Fig. S13). This mechanism aligns with the observed positive relationship between precipitation and LF incidence (46) and clarifies that recruitment is the process linking rainfall to infection peaks.

We further show that the seasonal pattern of LF outbreaks can be reproduced by predicted numbers of infected rodents (Fig. 4C), but not by population-level demographic drivers alone (SI Appendix, Fig. S14). This suggests that rodent infection dynamics, rather than demography *per se*, determine the timing of zoonotic hazard. The result is consistent with evidence that most human LF cases arise from direct contact with infected MN (22) and with theoretical work linking LF seasonality to rodent recruitment (47). Importantly, our approach demonstrates that data generalizability is feasible: by calibrating models using data from a Lassa-free but data-rich region, we can predict outbreak timing in data-sparse (2) but endemic areas using only climate inputs. Despite phylogeographic differences between LASV-carrying MN in Nigeria (sub-clade A1) and MORV-carrying MN in Tanzania (sub-clade B5) (21), similar demographic responses to climatic cues (31) support partial generalization between systems.

In contrast, the model did not predict outbreak magnitude (Fig. 4D), likely because outbreak size is influenced by human-related factors such as behavior, land use, case detection, and seasonal rodent movement into houses (27, 32, 46). These processes, while important, were beyond the scope of our analysis which focused on broad-scale demographic and climatic drivers.

A key limitation of our study is that we inferred LASV dynamics from parameters estimated on MORV, a non-pathogenic analogue. Differences between the viruses or host populations could bias our results, but validation is currently constrained by scarce LASV field data (2). The use of MORV and closely-related Mopeia virus as LASV surrogates is well established (24, 29, 30), yet future collection of MN/LASV field data would enable more direct calibration of our model. Agricultural practices, which influence MN movement and abundance (27), were also excluded due to data unavailability that would limit the generalizability of our model to other regions. However, such data could improve finer-scale models linking rodent ecology to human exposure (48). Such models could be calibrated on expanded serological datasets sampled from rodents and people inhabiting endemic regions. Our framework can be readily extended to other climate-sensitive rodent–pathogen systems, such as Sin Nombre virus in deer mice (17) or Puumala hantavirus in bank voles (18), or even other systems beyond rodent-borne diseases, providing a generalizable tool to study how environmental variation shapes zoonotic transmission.

In conclusion, our results disentangle the relative contributions of density-dependent and climatic factors to the population and infection dynamics of a key rodent reservoir. Beyond advancing understanding of MN ecology, this work demonstrates how long-term ecological data can inform mechanistic, transferable models of zoonotic risk. With expanded datasets from endemic regions, such models could underpin predictive tools to support LF preparedness and broader public health responses under a changing climate.

## Materials and Methods

### Rodent demographic and serological data

We extracted publicly available CMR data on MN from a 29-year study in Morogoro, Tanzania (6°51’S, 37°38’E) (28). Trapping occurred approximately every 28 days over three consecutive nights, with individual IDs, body weight, sex, and reproductive condition recorded. Trapping sessions were defined as periods separated by >3 days (28), yielding 377 sessions over 1,083 trap days. We excluded records missing key fields (e.g., ID, weight, reproductive condition), or with erroneous weight values (i.e., zero). Individuals were classified as sub-adults or adults based on reproductive condition (28). We calculated the Minimum Number Alive (MNA) as an index of relative abundance (49) and restricted analyses to females (*N*=8,628 individuals, 20,249 captures,1994–2023), under the assumption that male availability was not limiting for reproduction.

We obtained MORV seroprevalence data collected 2007–2017 using indirect immunofluorescence (34), excluding observations from 2007–2009 due to quality issues (J. Mariën, pers. comm.) and any records with missing/ambiguous serostatus. Seroprevalence was estimated per session for sub-adults and adults with 95% binomial confidence intervals. Because sex was not a significant predictor of exposure (multivariable generalized linear model (GLM), *P*=0.33), data were pooled across sexes, yielding 3,598 individuals and 7,850 tests across 79 sessions.

### Climate data

Daily precipitation and minimum/maximum temperatures were extracted at the trapping site (35, 36) and used to calculate mean temperature and 28-day rolling averages (right-aligned) for both variables, matching the temporal resolution of the trapping data. We applied seasonal-trend decomposition via locally estimated scatterplot smoothing (LOESS) (STL) (50) to extract trend, seasonal (year-independent), and residual (hereafter ‘variable’) components. LOESS windows were set to 5 years (trend) and periodic (seasonality). A robust fitting procedure and additional data cleaning steps were used to minimize the influence of anomalies (*SI Appendix*).

### Lassa fever case data

We used a web-scraping tool (51) to obtain weekly counts of laboratory-confirmed LF cases from the Nigerian Centre for Disease Control and Prevention (26). We extracted data for 2018–2025 due to improvements in testing capacity in early 2018 (52) and selected the five highest-burden states (Bauchi, Ebonyi, Edo, Ondo, and Taraba; *N*=6,469) (26), enabling predictions across different regional climates while limiting bias from including states with insufficient surveillance or low LF incidence. To reflect uncertainty in true case numbers, we sampled *N*=1,000 new Poisson-distributed case counts (with rate parameter *λ* set to reported counts). For each derived case count, we transformed dates of case reporting to dates of infection using an estimated LF incubation period posterior distribution (*SI Appendix*).

### Demographic process models

Bayesian hierarchical GLMs were developed in Stan (53) for body weight change (hereafter ‘growth’), recruitment, and survival, predicting demographic outcomes at time *t* + 1 based on individual traits at *t* and climate covariates at lagged time *t* − *l* (see below). These models informed an IPM (54) of population and infection dynamics. Full model details are in the *SI Appendix*.

Growth was estimated from paired captures of non-pregnant, non-lactating females and modelled with a heteroskedastic normal GLM where variance was a function of body weight. Recruitment was defined as non-pregnant, non-lactating individuals captured one session later as pregnant/lactating or two sessions later as lactating, consistent with known reproductive timing (55). Since survival was not directly observable from the data we modelled, using recapture histories, the probability of recapture and later adjusted these estimates for survival (*SI Appendix)*. Recruitment and recapture were modelled using GLMs with binomial error structures.

All models included MNA, body weight, and lagged climate predictors (temperature and precipitation) decomposed into trend, seasonal, and variable components. Climate lags were defined as 0–168 days in 28-day intervals (i.e., *l* = {0, 28, …, 168}), balancing ecological relevance and computational feasibility. Each *ith lag comb*ination was associated with a weight *ω* _*i*_ (where ∑*ω* = 1), allowing the model to learn optimal lag structures.

### Model fitting, validation, and selection

Models were fit using Stan’s No-U-Turn Sampler (NUTS) (53) via 4 chains of 2,000 iterations (1,000 burn-in) and convergence assessed using effective sample size (≥1,000) (56) and ensuring 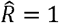 (53). Covariates were unit-scaled, and parameters given weakly informative prior distributions (zero-centered normal for *α* and *β*, symmetric Dirichlet for *ω*). For each demographic process, we fit 64 models representing all combinations of six climate predictors (2^6^). All included body weight and MNA. Model performance was evaluated using marginal log-likelihood via bridge sampling (57); top-ranked models were used for inference. Posterior predictive checks (58) were used to evaluate model fit to observed means and standard deviations, both overall and by month.

### Integral projection model

We developed an IPM (54) in R (59) to link demographic processes to rodent population and pathogen dynamics (https://github.com/gcmilne/lassa-popdyn; Fig. 1). The IPM tracks the distribution of body weights in the whole population and the pregnant subpopulation in discrete 28-day time steps (matching the temporal resolution of trapping data (28)), with transitions occurring according to time-specific probabilities of growth, survival, and pregnancy. Individuals are born as sub-adults with some weight distribution derived from data (60) and then mature into adults—who can then become pregnant—at a rate set to obtain a minimum age of first litter of 140 days (61). Individuals who give birth at time *t* can return to the pregnant sub-population at time *t* + 2, consistent with empirical data on MN breeding (61). Disease dynamics were modelled by nesting within the IPM a compartmental model: susceptible (*S*), horizontally infected (*H*), vertically infected (*V*), and recovered (*R*). We distinguished between horizontal and vertical transmission since studies on arenaviruses including MORV and LASV show that while the former generates a transient infection (with recovery rate *y; Fig. 1)*, vertical infections are chronic (24, 25, 44). Horizontal transmission is modelled with rate *β and vertica*l transmission from an infected pregnancy with probability φ. We use a mass-action model and assume a homogenously mixed population, consistent with MN’s non-territorial nature (62). Offspring born to recovered mothers receive maternal antibodies that offer transient protection against infection for 28 days (34, 63).

### Estimating transmission parameters

We estimated transmission parameters φ and *β* by comparing modelled sub-adult and adult seroprevalence to MORV seroprevalence data (34), withholding 2017 data for validation, using joint binomial log-likelihoods, with a model burn-in period of 5 years to allow dynamics to stabilize. Time-specific seroprevalence was modelled as [*N*(*t*) - *S*(*t*)]/*N*(*t*) reflecting life-long seropositivity after exposure (24). We employed a novel Bayesian adaptive grid search algorithm to approximate parameter posterior distributions, propagating uncertainty from demographic process inference into transmission parameter estimation. We defined prior distributions as φ ~ *Beta*(20,2) and *β* ~*Uniform*(1 × 10^−5^, 1.5 × 10^−3^) to reflect, respectfully, vertical transmission estimates from previous arenavirus studies (24, 31, 44, 64) and the absence of strong prior evidence on horizonal transmission. For *N*=1,000 demographic model posterior draws, the 2-dimensional parameter space was discretized into a 20×20 grid which was jittered with uniform noise, and the IPM simulated across this jittered grid. Unnormalized log-posterior densities were computed by summing joint binomial log-likelihoods and log-prior distributions. These were transformed into posterior probabilities via log-sum-exp transformation. Parameter combinations with weights below a threshold *τ* = 1 × 10^−7^ were discarded, and the remainder used to construct a finer grid spanning the high-density region of parameter space. For each demographic draw, this fine grid was re-evaluated to yield a smoother posterior probability surface *PD*_*i*_(φ, *β*).

### Posterior predictions

Posterior predictive simulations were generated using Bayesian melding (65). For *N*=1,000 iterations, a demographic posterior draw *i* was resampled with probability proportional to the marginal posterior mass across its *PD*_*i*_(φ, *β*). Conditional on that draw, a transmission parameter pair (φ, *β*) was then sampled from its grid with probability proportional to *PD*_*i*_(φ, *β*) and the IPM simulated. Posterior predictive medians and 95% CrI were summarized by taking the 0.5, 0.025, and 0.975 quantiles of simulations.

We assessed the consistency of 2017 hold-out seroprevalence data *θ*_*g*_(*t*) with predicted seroprevalence 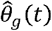 (computed as the median across *N* posterior simulations) using R package *brms* (56) to develop a Bayesian binomial GLM. This took the form: 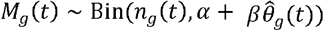, where *n*_*g*_(*t*) denotes number of rodents tested, *M*_*g*_(*t*) denotes number of seropositive rodents, *g* is group (sub-adult or adult), and *t* is time. The model was fit using 4 MCMC chains of 2,000 iterations. We compared this to an intercept-only null model by approximate leave-one-out cross-validation on expected log-predictive densities (56).

We recalibrated the model to assess its ability to capture the seasonal dynamics of human LF outbreaks in Nigeria (*SI Appendix*). Briefly, we extracted climate data (35, 36) at the centroid of each state, decomposed it as previously, and made posterior predictions of the number of rodents in compartments *H* and *V*, made relative to the maximum number over the time series. We compared predictions to the transformed LF cases (26) by calculating the difference between the timing and magnitude of predicted and observed peaks, for each state, year, and posterior simulation. We excluded from these estimates data from the 2020/2021 season due to interruptions in case reporting during the COVID-19 pandemic (26).

## Data Availability

The following datasets are publicly available: *M. natalensis* capture-mark-recapture data (https://www.nature.com/articles/s41597-023-02700-3), climate data (https://downloads.psl.noaa.gov/Datasets/), Lassa fever case data (https://ncdc.gov.ng/diseases/sitreps/?cat=5&name=An%20update%20of%20Lassa%20fever%20outbreak%20in%20Nigeria). Lassa fever incubation period data are given in the Supplementary Information file. The Morogoro arenavirus serological data (https://pubmed.ncbi.nlm.nih.gov/31545505/) are not open access; potential collaborators are encouraged to contact J. Marien

https://downloads.psl.noaa.gov/Datasets/

https://ncdc.gov.ng/diseases/sitreps/?cat=5&name=An%20update%20of%20Lassa%20fever%20outbreak%20in%20Nigeria

https://pubmed.ncbi.nlm.nih.gov/31545505/

## Acknowledgments

The authors acknowledge Research Computing at the James Hutton Institute for providing computational resources and technical support for the “UK’s Crop Diversity Bioinformatics HPC” (BBSRC grants BB/S019669/1 and BB/X019683/1), use of which has contributed to the results reported within this paper. GCM acknowledges support from a Sir Henry Dale Research Fellowship awarded to DWR, funded by the Wellcome Trust and the Royal Society (grant numbers 220179/Z/20/Z and 220179/A/20/Z). LAA acknowledges support from NERC (via QMEE CDT, grant number NE/P012345/1) and the Leverhulme Trust. JM, LK and HK acknowledge support from the Research Foundation - Flanders (FWO), grant number G065720N. CAD acknowledges support from the NIHR Health Protection Research Unit in Emerging and Zoonotic Infections and the Oxford Martin Programme in Digital Pandemic Preparedness. KEJ and DWR acknowledge support from the Trinity Challenge, Sentinel Forecasting Project. DWR also acknowledges funding from an Ecology and Evolution of Infectious Disease Award in collaboration with UK Research and Innovation (grant number 2208034), a Biotechnology and Biological Sciences Research Council Award (grant number BB/X005364), and the NSF EEID (grant number 2109828).

