## Supplementary material for "Climate-driven rodent infection dynamics align with Lassa fever seasonality in humans": SI Appendix

PNAS

##### This PDF file includes:

Supporting text
Figs. S1 to S14
Tables S1 to S3
SI References

### Supporting Information Text

#### 1. Supplementary methods

**A. Climate data.** Daily precipitation and temperature data (1, 2) were extracted at the trapping site (3). To account for erroneous temperature data, we compared each daily value to a long-term mean (1991–2020) (4). Missing daily values and daily values with $\geq$60% absolute difference from the long-term mean were interpolated using neighbouring daily values. 28-day rolling averages were calculated for precipitation and temperature separately, matching the temporal resolution of the trapping data (3). These were subsequently decomposed into trend, seasonal, and residual (hereafter 'variable') components using seasonal-trend decomposition via locally estimated scatterplot smoothing (LOESS) (STL) (5), with a five-year trend window and a periodic seasonal window.

**B. Statistical models.** Bayesian hierarchical generalised linear models (GLMs) were formulated using long-term capture mark recapture data (3) to describe the demographic processes of body weight change, pregnancy, and survival. The vector of covariates, $\boldsymbol{X}$, for each model were: weight ($w$), population size ($N(t)$), temperature trend ($t_{trend}$), temperature seasonality ($t_{seas}$), temperature variability ($t_{var}$), precipitation trend ($p_{trend}$), precipitation seasonality ($p_{seas}$), and precipitation variability ($p_{var}$),

$$\boldsymbol{X}(t) = \{w, N(t), t_{\text{trend}}(t), t_{\text{seas}}(t), t_{\text{var}}(t), p_{\text{trend}}(t), p_{\text{seas}}(t), p_{\text{var}}(t)\} \qquad [1]$$

with corresponding vector of coefficients

$$\boldsymbol{\beta} = \{\beta_w, \beta_N, \beta_{t_{\text{trend}}}, \beta_{t_{\text{seas}}}, \beta_{t_{\text{var}}}, \beta_{p_{\text{trend}}}, \beta_{p_{\text{seas}}}, \beta_{p_{\text{var}}}\}. \qquad [2]$$

Labelling the elements of the vectors of covariates and coefficients as $\boldsymbol{X} = \{X_1, X_2, ..., X_8\}$ and $\boldsymbol{\beta} = \{\beta_1, \beta_2, ..., \beta_8\}$, the demographic process models took the form

$$g(\mu) = a + \beta_1 X_1 + \beta_2 X_2(t) + \sum_{i=3}^{8} \beta_i \sum_{k=1}^{K} \omega_k X_i(t - L_{i,k}) \qquad [3]$$

where $g(\mu)$ is the link function (logit($p$) for the Bernoulli processes of survival and pregnancy, and $\mu$ for the mean of the continuous process of body weight change), $L_{i,k}$ is the $k$th temperature/precipitation lag corresponding to covariate $i$, and $\omega_k$ is that corresponding lag combination's assigned weight (see Section B.1).

The effect of body size on individual-level recruitment could only be observed in female rodents (via observable pregnancy and lactation). Therefore, body growth, pregnancy, and survival were only modelled directly in female rodents, with a separate model fit to estimate corresponding male population size. Coefficients for each demographic process are indicated with subscripts: $B$ for body weight change, $S$ for survival, $P$ for pregnancy, and $M$ for male population size (see Section B.5).

***B.1. Climate lags.*** We introduced lags to the decomposed climate data ($X_3, ..., X_8$ in Eq. 1) (1, 2). We defined temperature lags $l_T = \{0, 28, ..., 168\}$ and precipitation lags $l_P = \{0, 28, ..., 168\}$ (units in days), giving $K = |l_T||l_P| = 7 \times 7$ lag combinations for each demographic process model. Lag weights, $\omega$, associated with each lag combination were defined as a $(K - 1)$ simplex, where $\omega_k \geq 0$ for $k \in \{1, ..., K\}$ and $\sum_{k=1}^{K} \omega_k = 1$. A uniform Dirichlet prior distribution (with all concentration parameters equal to one) was placed on this simplex ($\boldsymbol{\omega} \sim \text{Dir}(\boldsymbol{\alpha} = \{1, 1, ..., 1\})$). Lag weights $\boldsymbol{\omega}$ were estimated separately for each model, contributing to model likelihood through a weighted sum (Eq. 3). In Eq. 3, $L_{i,k} =$ takes the value of one of the $\{l_T, l_P\}$ pair corresponding to the $k$th combination of temperature-precipitation lags. If $i$ corresponds to a temperature covariate ($i \in \{3, 4, 5\}$), then $L_{i,k}$ takes the value of the temperature lag, while if $i$ corresponds to a precipitation covariate ($i \in \{6, 7, 8\}$) then $L_{i,k}$ is the paired precipitation lag.

***B.2. Body weight change.*** Changes in body weight were observed by identifying consecutive individual capture records and recording the weight at all captures. Subsequent trapping sessions were defined as 3–5 weeks after the initial capture to allow for variability from the 28-day mean time between trapping sessions (3). An individual's body weight $Y$ at time $t + 1$ given it had a body weight $w$ at time $t$ was modelled as a heteroskedastic normal distribution

$$Y \sim \text{N}(\mu_B(w, t), \sigma_B(w)), \quad \text{where} \quad \sigma_B(w) = \exp(\alpha_\sigma + \beta_\sigma w), \qquad [4]$$

with the mean of the distribution $\mu_B$ described as in Eq. 3 ($g(\mu_B) = \mu_B$), noting that the individual's body weight at time $t$ is the first covariate in the vector of covariates ($w = X_1$). The probability density function of $Y$ is then given by

$$p_B(y|w, t) = \frac{1}{\sigma_B(w)\sqrt{2\pi}} \exp\left(-\frac{1}{2}\left(\frac{v - \mu_B(w, t)}{\sigma_B(w)}\right)^2\right). \qquad [5]$$

***B.3. Survival.*** Survival, whether an individual of body weight $w$ at time $t$ survives to time $t+1$, was modelled as a random variable drawn from a Bernoulli distribution with probability of success (survival) $p_S$. Since survival could not be directly observed from the data, we first modelled the probability of recapture, $p_R$, using the model outlined in Eq. 3 with a logit-link function for $g(\mu)$. We then related $p_S$ to $p_R$ by conditioning on whether the individual is recaptured or not:

$$\begin{aligned} p_S(w,t) &= \mathbb{P}(\text{alive at } t+1) \\ &= p_R(w,t) + \mathbb{P}(\text{alive at } t+1 | \text{ never recaptured after } t) \cdot (1 - p_R(w,t)). \end{aligned} \qquad [6]$$

Since $\mathbb{P}(\text{alive at } t+1 | \text{ never recaptured after } t)$ could not be known from the data, we defined this as $\kappa(w,t)$: the probability that an individual captured at time $t$ of weight $w$ is still alive at time $t+1$ despite never being recaptured (allowing for factors such as migration and stochasticity). It then follows that

$$p_S(w,t) = \kappa(w,t) + (1 - \kappa(w,t))p_R(w,t). \qquad [7]$$

Since it would not be possible to distinguish between $\kappa$ and $p_R$ if $\kappa$ were allowed to vary by weight and time, $\kappa$ was assumed constant (i.e., $\kappa(w,t) = \kappa$) and calibrated to obtain stable population dynamics (see Section F.4).

***B.4. Pregnancy.*** We modelled pregnancy as the probability of a (not pregnant) female of weight $w$ at time $t$ being pregnant at time $t+1$. While recent pregnancy was observable from the data (3), inclusion of these individuals would bias estimates of the effect of weight on the probability of pregnancy. To avoid this, the data were restricted to females who were not pregnant at time $t$ but were pregnant or lactating when captured in the next trapping session at time $t+1$ (3–5 weeks after initial capture), or lactating but not pregnant when captured in the subsequent trapping session at time $t+2$ (6–10 weeks after initial capture). This strategy was informed by the known reproductive timings of *M. natalensis*, of which the average gestation period is approximately 21 days, with visible pregnancy apparent in the second half of this period, followed by lactation for approximately 21 days (6). Similarly to survival, pregnancy was modelled using a Bernoulli likelihood and a logit-link function (Eq. 3).

***B.5. Male population size.*** The integral projection model (IPM) models the female portion of the population (see Section F). However, since the demographic process models were fit using total population size (males and females) as a covariate ($X_2$ in Eq. 1), it was necessary to estimate total population size to ensure correct calibration of density dependence in the IPM.

To achieve this, we formulated and compared Bayesian GLMs to estimate male population size, $N_m$, as a function of the observed, time-specific female population size, $N_f(t)$, using a zero-truncated normal distribution (see Section D for model details). The best-fitting model (see Section 2) was of the form:

$$N_m \sim \text{TN}(\mu_M, \sigma_M^2, 0, \infty), \quad \text{where} \quad \mu_M = \alpha_M + \beta_M N_f(t) + u_M. \qquad [8]$$

Here, $\alpha_M$ is the global intercept, $\beta_M$ is the effect of female population size, and $u_M$ represents the random intercept for month (included to capture the time-dependent relationship between male and female population sizes). The standard deviation $\sigma_M$ captures residual variation in the male population size. The distribution is truncated at zero to ensure no non-negative values for number of males $N_m$. By estimating this relationship, the IPM could dynamically update total population size at each time step, maintaining accurate density dependence (see Section F).

**C. Fitting statistical models.** The four statistical models of body weight change, survival, pregnancy, and male population size were fit in Stan (7) using a No-U-Turn Sampler (NUTS) (8) Hamiltonian Monte Carlo algorithm, with prior distributions as specified in Table S1. The observations $x_j$ of a given covariate were transformed to unit scale (mean of 0, standard deviation of 1) by $\hat{x}_j = \frac{x_j - \bar{x}_j}{\sigma_{x_j}}$, where $\bar{x}_j$ represents the mean across measured values of $x_j$ and $\sigma_{x_j}$ the standard deviation across measured values of $x_j$. For models of survival and pregnancy we estimated the coefficients $\alpha$ and $\boldsymbol{\beta}$, and lag weights $\boldsymbol{\omega}$ (Eq. 3), while for body weight change we estimated these parameters plus $\alpha_\sigma$ and $\beta_\sigma$ (Eq. 4; Table S1). Since $\boldsymbol{\omega}$ is comprised of $K$ elements (denoting the total number of lag combinations; Section B.1), model estimation provided $K$ posterior distributions for $\boldsymbol{\omega}$ for each demographic process. Hence, we estimated $K+9$ parameters for survival and pregnancy and $K+11$ for body weight change. For male population size, we estimated four parameters ($\alpha_M$, $\beta_M$, $u_M$, and $\sigma_M^2$; Eq. 8). Models were fit to their respective datasets using four independent Markov chain Monte Carlo (MCMC) chains run for 2,000 iterations each, discarding the first 50% of iterations as burn-in. Convergence was assessed by ensuring $\hat{R} = 1$ (7) and effective sample size $\hat{N} \geq 1000$ (9), and by visually inspecting MCMC trace plots. Correlation between posterior parameter values was assessed via cross-correlation plots.

**D. Model comparison.** For each demographic process (body weight change, survival, pregnancy), we fit models containing every possible combination of the six climate predictors ($X_3, .., X_8$ in Eq. 3), giving $2^6 = 64$ models for each demographic process. For each model $M_i$ ($i \in \{1, 2, ...64\}$) we then derived via bridge sampling (10) an approximation of the marginal log-likelihood,

$$p(y|M_i) = \int p(y|\theta, M_i)p(\theta|M_i)d\theta \qquad [9]$$

where $p(\boldsymbol{y}|\theta, M_i)$ is the likelihood of the parameters $\theta$ in the model $M_i$ (given observations $\boldsymbol{y}$), and $p(\theta|M_m)$ is the prior distribution. The relative evidence for one model over another was then computed as a Bayes factor,

$$F(M_i, M_j) = \frac{p(\boldsymbol{y}|M_i)}{p(\boldsymbol{y}|M_j)}. \quad [10]$$

For male population size (see Section B.5), we estimated four models of varying complexity using the R package *brms* (9). All models contained female population size as a coefficient. Model 1 contained a fixed intercept. Model 2 contained a random intercept for month of sampling. Model 3 had a fixed intercept and a random coefficient for month. Model 4 had a random intercept plus a random coefficient for month. After model fitting, we made pairwise comparisons of each model's predictive accuracy using approximate leave-one-out cross-validation (LOO-CV) on expected log-predictive densities (ELPD)(9), with higher ELPD indicating better performance. If there was no difference in the ELPDs of two competing models, the simpler model was preferred in line with the principle of parsimony.

**E. Posterior predictive checks.** Posterior predictive checks (PPCs) were performed to assess the ability of each demographic model to capture relevant aspects of the data (e.g., mean, standard deviation). For each model, data for PPCs were generated by simulating from the posterior predictive distribution. The posterior predictive distribution for replications $\boldsymbol{y}_{\text{rep}}$ of the observed data $\boldsymbol{y}$ given model parameters $\theta$ can be expressed as

$$p(\boldsymbol{y}_{\text{rep}}|y) = \int p(\boldsymbol{y}_{\text{rep}}|\theta)p(\theta|\boldsymbol{y})d\theta. \quad [11]$$

Note that the notation $\boldsymbol{y}_{\text{rep}}$ indicates that the same predictors $\boldsymbol{X}$ are used to simulate from the posterior predictive distribution as are used to fit the model.

#### F. Integral projection model.

***F.1. Model structure.*** We constructed an age- and weight-structured IPM to simulate population and infection dynamics of *M. natalensis* over discrete time steps $t = \{0, 1, ..., t_{\max}\}$ (with each time step representing 28 days to match the data's temporal resolution (3)), based on the fitted demographic process models of body weight change, survival, and pregnancy (Section B). Model parameters are given in Table S1.

We denote the density of individuals of weight $w$ and age $a$ in the population at time $t$ as $D(w, t, a)$, where $a \in 1, 2$ denotes the sub-adult class and $a \geq 3$ denotes the adult class. The density of pregnant individuals is denoted $J(w, t)$ (so note that $J(w, t) \leq \sum_{a \geq 3} D(w, t, a)$, since all pregnant individuals are adults). Individuals at time $t$ determined to become pregnant (with probability $p_P(w, t)$; Section B.4) move to compartment $J$ at time $t + 1$. It was then assumed that if the pregnant individual survives to $t + 2$, they recruit $l$ female offspring where $l$ is a constant (set to 5 (3)). Offspring follow a body weight distribution given by $v(w)$ which was parameterised with data (11). Consistent with evidence showing that female *M. natalensis* can become pregnant approximately 4 weeks after giving birth (12), the model allows individuals in compartment $J$ at time $t$ to leave the compartment at time $t+2$. Individuals are born into the first sub-adult class ($a = 1$) and reach the adult class ($a = 3$) at three time steps (i.e, 84 days), meaning they can then become pregnant and can give birth to offspring at the fifth time step (140 days), approximately matching average age of first litter estimates from empirical data (130 days (12)). For ease, we will define $D(w, t)$ as the marginalised density of body weight $w$ at time $t$ across both age classes (i.e., $D(w, t) = \sum_a D(w, t, a)$). Noting the pregnancy probability $p_P$ is only applied to the adult population ($a \geq 3$) and that sub-adults ($a = 1, 2$) are governed by the same growth and survival functions as adults (conditioned on weight), the model is then given by the integral projection equations:

$$\begin{aligned} J(w, t+1) &= \int_0^\infty p_B(w|y, t)p_S(y, t)p_P(y, t)\left(\sum_{a \geq 3} D(y, t, a) - J(y, t)\right) dy \\ D(w, t+1, 0) &= \int_0^\infty l p_S J(y, t)v(w)\, dy \\ D(w, t+1, a+1) &= \int_0^\infty p_B(w|y, t)p_S(y, t)D(y, t, a)\, dy \quad \text{for } a \geq 1 \end{aligned} \quad [12]$$

To implement the model computationally, Eq. 12 was discretised by converting probability density functions, $p_G(w, t)$ and $v(w)$, into probability mass functions and converting integrals into sums over a set of possible weights $W = \{1, 2, 3, ..., 100\}$ (in grams; chosen to capture the full possible range of weights since observed rodents did not exceed 96 grams (3)), giving the discretised form of the model:

$$
\begin{aligned}
J(w,t+1) &= \sum_{y\in W} p_B(w|y,t)p_S(y,t)p_P(y,t)\left(\sum_{a\geq 3} D(y,t,a) - J(y,t)\right) \\
D(w,t+1,0) &= \sum_{y\in W} lp_S(y,t)J(y,t)v(w) \\
D(w,t+1,a+1) &= \sum_{y\in W} p_B(w|y,t)p_S(y,t)D(y,t,a) \quad \text{for } a\geq 1
\end{aligned} \tag{13}
$$

where now the compartments $D(w,t,a)$ and $J(w,t)$ refer to the number, rather than density, of individuals of weight $w$ at time $t$. Note that all adult age classes ($a \geq 3$) are treated equally and therefore the system can be implemented with just three age classes ($a = 1$, $a = 2$, and $a \geq 3$) rather than tracking ages of adults.

***F.2. Infection dynamics.*** The IPM was expanded to include pathogen transmission dynamics in a compartmental model of the form: susceptible (S), horizontally infected (H), vertically infected (V), and recovered (R) (see main text figure). This structure was motivated by empirical data and previous models of arenavirus dynamics wherein vertically infected individuals remain chronically infectious for life whereas horizontally infected individuals are transiently infectious and recover in approximately 30 days (13). The model takes the form,

$$
\begin{aligned}
\frac{dS}{dt} &= b(S+(1-\varphi)(H+V)) + \eta R - \beta(t)S(H+V) - mS \\
\frac{dH}{dt} &= \beta(t)S(H+V) - \gamma H - mH \\
\frac{dV}{dt} &= \varphi b(H+V) - mV \\
\frac{dR}{dt} &= bR + \gamma H - \eta R - mR
\end{aligned} \tag{14}
$$

where $\beta(t)$ is the horizontal transmission rate (estimated; Section G), $\gamma$ is the recovery rate following horizontal transmission ($28^{-1}$ days (13)), $m$ is the intrinsic mortality rate, $b$ is the intrinsic birth rate, $\varphi$ is the probability of vertical transmission between an infected individual and its offspring (estimated; Section G), and $\eta$ is the rate of loss of transient maternal antibodies acquired from an offspring birthed by a recovered mother ($28^{-1}$ days (14)) (Table S1).

To nest this compartmental model within the IPM, it was first necessary to estimate the number of horizontal infections between time steps. We estimated the horizontal force of infection, $\lambda(t)$, using a fourth-order Runge-Kutta (RK4) method as follows. At time step $t$, the susceptible ($S_t$), horizontally-infected ($H_t$), and vertically-infected ($V_t$) populations are defined by integrating over their respective weight distributions (e.g., $H_t = \int_0^\infty D_H(w,t)dw$). By assuming no demographic or infection changes between time steps, the number of new horizontal infections between time $t$ and $t+\tau$ (where $\tau \in (0,1]$), $K_t(\tau)$, can be shown to follow a susceptible-infected (SI) version of Eq. 14:

$$
\begin{aligned}
\frac{dS}{d\tau} &= -\beta SI, \\
\frac{dI}{d\tau} &= \beta SI, \\
\frac{dK_n}{d\tau} &= \beta SI,
\end{aligned} \tag{15}
$$

where $S(\tau=0) = S_t$, $I(\tau=0) = H_t + V_t$, and $K_t(\tau=0) = 0$. Using RK4, we estimate $K_t(\tau=1)$ with the following slopes:

$$
\begin{aligned}
k_1 &= \beta(t)S_tI_t, \\
k_2 &= \beta(t)\left(S_t - \frac{k_1}{2}\right)\left(I_t + \frac{k_1}{2}\right), \\
k_3 &= \beta(t)\left(S_t - \frac{k_2}{2}\right)\left(I_t + \frac{k_2}{2}\right), \\
k_4 &= \beta(t)(S_t - k_3)(I_t + k_3).
\end{aligned} \tag{16}
$$

The total number of new infections over the time step is

$$
E_t = K_t(1) = \frac{1}{6}(k_1 + 2k_2 + 2k_3 + k_4). \tag{17}
$$

The force of infection on each susceptible individual (assumed to be independent of body weight) is then given by

$$\lambda(t) = \frac{E_t}{S_t} \tag{18}$$

The compartmental model is then nested within the (discretised) IPM:

$$\begin{aligned}
B_S(t+1) &= l \sum_{y \in W} p_S(y,t) J_S(y,t) \\
B_I(t+1) &= l \sum_{y \in W} p_S(y,t) J_{H+V}(y,t) \\
B_R(t+1) &= l \sum_{y \in W} p_S(y,t) J_R(y,t)
\end{aligned} \tag{19}$$

$$\begin{aligned}
D_S(w,t+1,1) &= \big(B_S(t+1) + (1-\varphi) B_I(t+1)\big) v(w) \\
D_H(w,t+1,1) &= 0 \\
D_V(w,t+1,1) &= \varphi B_I(t+1) v(w) \\
D_R(w,t+1,1) &= B_R(t+1) v(w)
\end{aligned} \tag{20}$$

$$\left.\begin{aligned}
D_S(w,t+1,a+1) &= \textstyle\sum_{y \in W} p_B(w|y,t) p_S(y,t) \big((1-\lambda(t)) D_S(y,t,a) + \chi_{\{1\}}(a) D_R(y,t,a)\big) \\
D_H(w,t+1,a+1) &= \textstyle\sum_{y \in W} p_B(w|y,t) p_S(y,t) \lambda(t) \sum_{a \geq 3} D_S(y,t,a) \\
D_V(w,t+1,a+1) &= \textstyle\sum_{y \in W} p_B(w|y,t) p_S(y,t) D_V(y,t,a) \\
D_R(w,t+1,a+1) &= \textstyle\sum_{y \in W} p_B(w|y,t) p_S(y,t) \big((1-\chi_{\{1\}}(a)) D_R(y,t,a) + D_H(y,t,a)\big)
\end{aligned}\right\} \text{for } a \geq 1 \tag{21}$$

$$\begin{aligned}
J_S(w,t+1) &= \sum_{y \in W} p_P(y,t) \big(1-\lambda(t)\big) p_B(w|y,t) p_S(y,t) \sum_{a \geq 3} D_S(y,t,a) \\
J_{H+V}(w,t+1) &= \sum_{y \in W} p_P(y,t) p_B(w|y,t) p_S(y,t) \sum_{a \geq 3} \big(\lambda(t) D_S(y,t,a) + D_V(y,t,a)\big) \\
J_R(w,t+1) &= \sum_{y \in W} p_P(y,t) \Big(p_B(w|y,t) p_S(y,t) \sum_{a \geq 3} D_R(y,t,a) + p_B(w|y,t) p_S(y,t) \sum_{a \geq 3} D_H(y,t,a)\Big).
\end{aligned} \tag{22}$$

In this system of equations, $B_S$, $B_I$, and $B_R$ are the number of susceptible, infected, and recovered (maternally-antibody positive) juveniles, respectively, entering the population, and subscripts for $D$ and $J$ denote infection status within the whole population and the pregnant sub-population, respectively. $\chi_{\{1\}}(a)$ denotes the indicator function which takes the value of 1 if $a = 1$ (which here causes movement of sub-adults with maternal antibodies moving from recovered to susceptible infection status) and 0 otherwise. Modelled (female) population size can then be given by

$$N_f(t) = \sum_{w \in W} \sum_{a} \big(D_S(w,t,a) + D_H(w,t,a) + D_V(w,t,a) + D_R(w,t,a)\big) \tag{23}$$

***F.3. Calibrating density dependence.*** To ensure proper calibration of density dependence while simulating the IPM using Eqs. (19) to (22), $N_f(t)$ (Eq. 23) had to be adjusted to total population size. This ensured no discrepancy between the total population size data used to fit demographic process models ($X_2$ in Eqs. 1 and 3) and the population covariates used as inputs to fitted demographic process models during IPM simulation. At every model $t$, $N_f(t)$ was used to derive an estimated total population size, $N_{m,f}(t)$, by

$$N_{m,f}(t) = N_f(t) + N_m(t). \tag{24}$$

where $N_m$ was predicted at each time step of the IPM using Eq. 8 (using fitted parameter values for $\alpha_M$, $\beta_M$, and $\sigma_M$). Before input into fitted demographic process models (Eq. 3), $N_{m,f}(t)$ was further adjusted to bring the absolute modelled population size in line with the mean observed population size,

$$N(t) = \left(\frac{\bar{N}}{2N_0}\right) N_{m,f}(t) \tag{25}$$

where $\bar{N}$ is the mean observed Minimum Number Alive (male and female) (3) and $N_0$ is the initial modelled female population size (at $t = 0$). The term $2N_0$ accounts for the (assumed 1:1) sex ratio which was necessary to include because the demographic process models were fit to data on total population size (Section B) whereas the IPM only explicitly considered females but requires total population size as an input (Eqs. 8, 24, 25).

**F.4. Calibrating survival.** We calibrated $\kappa$ to obtain the probability of survival, $p_S$, from the probability of recapture, $p_R$ (Section B.3, Eqs. 6, 7), as follows. The IPM was simulated for $N = 1000$ random posterior draws of the fitted statistical model parameters (Section C) to obtain predictions of the time-specific modelled population size, $N(t)$ (Eq. 23). For each joint posterior draw, the IPM was simulated and values of $\kappa$ interpolated to obtain a modelled population size that was within 1% of the initial population size at $t = 0$, $N_0$. In subsequent model fitting (Section G) and posterior prediction (Section H), $\kappa$ was set to the median of this estimated distribution which ensured stable population dynamics.

**G. Fitting transmission parameters.** A Bayesian adaptive grid search algorithm was developed to approximate the posterior distributions of transmission parameters $\varphi$ and $\beta$, by fitting to Morogoro arenavirus seroprevalence data collected 2010–2016 (withholding 2017 data for validation) (14). We set prior distributions $\varphi \sim Beta(20, 2)$ (15, 16) and $\beta \sim \mathrm{U}(0.00001, 0.0015)$. The parameter space was discretised into a $20 \times 20$ coarse grid $\mathbf{C}$ over the intervals $\varphi \in [0.01, 0.99]$, $\beta \in [0.0001, 0.0015]$. For $i = \{1, 2, ..., N\}$ (where $N = 1000$) random independent draws from the demographic model posterior distributions, this coarse grid was adjusted by some random uniform noise $\varepsilon_i$ in both dimensions to obtain new grid values $\mathbf{C}^{(i)} = \mathbf{C} + \varepsilon_i$, where $\varepsilon_i$ for each parameter was uniform on $(-\frac{d}{2}, +\frac{d}{2})$, where $d$ is the (equal) difference between consecutive parameter values in the grid $\mathbf{C}$. The IPM was simulated using the sampled demographic parameter values for each of the coarse grid infection parameter values to derive modelled seroprevalences for sub-adults and adults by $\big(N(t) - S(t)\big)/N(t)$. These modelled seroprevalence outputs were compared to empirical data using joint binomial log-likelihood distributions, which were combined with log-prior distributions to obtain unnormalised log-posterior densities. Log-sum-exp transformation was used to transform unnormalised log-posterior densities into importance sampling weights. Parameter combinations with importance weights below a threshold $\tau < 1 \times 10^{-7}$ were discarded and the remainder used to construct a finer grid spanning this high density region of parameter space. For each demographic draw, this fine grid was re-evaluated to obtain a smoother posterior probability surface which was used to generate posterior predictions (see Section H).

**H. Posterior predictions.** To propagate uncertainty from previously estimated model parameters into downstream inference, we adopted a Bayesian melding framework (17, 18). Specifically, we first obtained posterior samples from four upstream statistical models (denoted collectively as $\Phi$; Section C) by fitting to demographic and climate data, $\mathcal{D}_1$. We then sought to estimate two additional transmission dynamics parameters, denoted collectively as $\phi$, by combining information from these upstream models with a second data source, $\mathcal{D}_2$, comprising seroprevalence observations (Section G).

For each upstream posterior sample $\Phi^{(i)} \sim p(\Phi|\mathcal{D}_1)$, we evaluated the conditional posterior distribution over $\phi$ given the seroprevalence data:

$$p(\phi|\Phi^{(i)}, \mathcal{D}_2) \propto p(\phi) \times \mathcal{L}_2(\mathcal{D}_2|\Phi^{(i)}, \phi), \qquad [26]$$

where $\mathcal{L}_2$ is the likelihood of the seroprevalence data under model parameters $(\Phi^{(i)}, \phi)$, and $p(\phi)$ is the prior distribution over the transmission parameters. This conditional posterior distribution was approximated over a 2D grid for each $\Phi^{(i)}$ (see Section G) to obtain a posterior density function $PD_i(\phi) = p(\phi|\Phi^{(i)}, \mathcal{D}_2)$. The joint posterior distribution over both upstream and transmission parameters can then be approximated by:

$$p(\Phi^{(i)}, \phi^{(j)}) \approx p(\Phi^{(i)}|\mathcal{D}_1) \cdot p(\phi^{(j)}|\Phi^{(i)}, \mathcal{D}_2). \qquad [27]$$

To sample efficiently from this joint distribution, we used importance resampling at two levels. First, we re-weighted the upstream samples $\Phi^{(i)}$ based on the concordance between modelled and empirical seroprevalence. Specifically, we marginalised over each conditional posterior $PD_i(\phi)$ to compute an integrated weight:

$$w_i \propto \int p(\phi|\Phi^{(i)}, \mathcal{D}_2)\, d\phi \approx \sum_j PD_i(\phi^{(j)}), \qquad [28]$$

where $PD_i(\phi^{(j)})$ are unnormalised posterior densities. To ensure numerical stability, we computed this sum on the log scale using log-sum-exp transformation:

$$\log w_i = \log \sum_j \exp\left(\log PD_i(\phi^{(j)}) - \max_j \log PD_i(\phi^{(j)})\right) + \max_j \log PD_i(\phi^{(j)}), \qquad [29]$$

These weights $w_i$ were then exponentiated and normalised to sum to 1. The resulting normalised weights reflect the compatibility of each upstream draw $\Phi^{(i)}$ with the transmission dynamics implied by the seroprevalence data $\mathcal{D}_2$. Second, for each selected draw $\Phi^{(i)}$, we generated samples of the transmission parameters $\phi$ by resampling from the corresponding conditional posterior surface $PD_i(\phi)$. Specifically, we treated the unnormalised log posterior densities $\log PD_i(\phi^{(j)})$ over the grid as importance weights, stabilised them using a log-sum-exp transformation, and normalised them to define a discrete probability distribution over $\phi$. This ensures that transmission parameter values are sampled with probability proportional to their posterior support, conditional on the selected upstream model $\Phi^{(i)}$. The Bayesian melding procedure thus proceeds as follows:

- For each $\Phi^{(i)}$, compute the conditional posterior surface $PD_i(\phi)$ over a grid.
- Marginalise over each $PD_i(\phi)$ to obtain an importance weight $w_i$ for $\Phi^{(i)}$ (Eq. 29).

- Draw $N = 1000$ samples of $(\Phi, \phi)$ by:
    1. Sampling $\Phi^{(i)}$ with probability proportional to $w_i$.
    2. Sampling $\phi^{(j)}$ from the discretised grid conditional on $\Phi^{(i)}$, with probability proportional to $PD_i(\phi^{(j)})$, i.e., the normalised posterior density for $\phi$ conditional on the selected $\Phi^{(i)}$.

Posterior predictive simulations were summarised by medians and 95% credible intervals (0.5, 0.025, and 0.975 quantiles).

#### I. Model recalibration for Nigeria.

***I.1. Posterior predictions.*** We re-calibrated the fitted model to Nigeria to determine the extent to which the seasonal patterns of Lassa fever outbreaks could be captured. Specifically, we focussed on the five Nigerian states with the highest burden of Lassa fever (Bauchi, Ebonyi, Edo, Ondo, and Taraba) (19) and extracted data for 2018–2025 due to improvements in testing capacity in early 2018 (20). We extracted daily temperature and precipitation data (1, 2) at the centroid of each state, decomposed these as previously (Section B), and for each state recalibrated survival adjustments ($\kappa$) using the state-specific climate data (Section F.4). We then made posterior predictions of infection dynamics for each state by simulating the model with the $N = 1000$ $(\Phi, \phi)$ draws (Section H), substituting the Morogoro climate data for the Nigerian state-specific climate data. For each Nigerian state $s$, we predicted the time-specific zoonotic hazard $H$ as the number of infected rodents:

$$H_s(t) = \sum_w \Big( D_{H,s}(w,t) + D_{V,s}(w,t) \Big) \qquad [30]$$

which we transformed into a relative metric by

$$H^*_s(t) = \frac{H_s(t)}{\max_t(H_s(t))} \qquad [31]$$

***I.2. Lassa fever case data.*** We compared predictions to weekly reports of confirmed Lassa fever cases from the Nigeria Centre for Disease Control and Prevention (19). Let $Y_s(d)$ be the reported cases in state $s$ on day $d$ and let us assume, due to imperfect sampling, that the reported case numbers are not the true cases numbers. We can then estimate uncertainty in the true case numbers by:

$$\hat{Y}_s(d) \sim \text{Poisson}(\lambda = Y_s(d)) \qquad [32]$$

To make model estimates comparable to the data, case data were transformed to obtain an infection time series by sampling from an inferred Lassa fever incubation period distribution $p_I$:

$$I_s(d) = \sum_{i=1}^{30} \hat{Y}_s(d+i) p_I(i) \qquad [33]$$

To make observed case data comparable to model predictions, this was transformed into a relative metric by:

$$I^*_s(d) = \frac{I_s(d)}{\max_d(I_s(d))} \qquad [34]$$

$p_I$ was inferred by collating published data on nosocomial infections of Lassa fever. Although infections in a healthcare setting are caused by human-to-human transmission rather than zoonotic transmission—and thus one might expect differences in virulence or viral dose that could lead to different incubation periods—we emphasise that we used the only available data to make these estimates. Papers were considered if they appeared in a PubMed or Scopus search carried out in September 2025 for "("Lassa fever") AND ("nosocomial" OR "hospital infection")". Individual cases of Lassa fever were included if they:

- were confirmed through laboratory testing
- were believed to have been infected in a healthcare setting
- had a finite and informative period for exposure (time window of 0–28 days)

For each included case ($N = 30$) we extracted data on study location, earliest and latest possible date of exposure, earliest and latest possible date of symptom onset, and fatality (including date of fatality if recorded) (Table S2).

We used the Metropolis-Hastings (MCMC) algorithm to fit a log-normal distribution (Lognormal$(\mu, \sigma^2)$) to the observed incubation period windows with an uninformative prior distribution. We took into account that the case data recorded possible windows of symptom onset rather than exact incubation periods by calculating likelihood as the probability of observing a set of incubation periods within the observed windows:

$$\ell(\mu, \sigma^2) = \prod_i \int_{w_i^- - \frac{1}{2}}^{w_i^+ + \frac{1}{2}} p(x; \mu, \sigma^2)\, dx = \prod_i \left( \text{erf}\left( \frac{\log(w_i^+ + \frac{1}{2}) - \mu}{\sigma} \right) - \text{erf}\left( \frac{\log(w_i^- - \frac{1}{2}) - \mu}{\sigma} \right) \right) \qquad [35]$$

where $p$ denotes the probability density function of the log-normal distribution, $i$ indexes the collated nosocomial cases, and $w_i^-$ and $w_i^+$ denote the minimum and maximum (respectively) possible incubation period (in days) based on the hospital observations recorded. The addition and subtraction of $\frac{1}{2}$ is to account for the log-normal distribution being continuous while records are in discrete numbers of days.

#### 2. Supplementary results

The best-performing model for growth (body weight change), as indicated by log marginal likelihoods, included coefficients for precipitation seasonality and precipitation trend. These coefficients were also present in the top 20.3% (13/64) of best-performing growth models (Figure S1). The best-performing and converging model for survival included precipitation seasonality and temperature trend. These coefficients were also present in the top 25% (16/64) of best-performing survival models (Figure S2). All 16 survival models including temperature seasonality did not converge (Figure S2), indicating the unsuitability of this variable for explaining survival. The best performing model for pregnancy included precipitation seasonality and precipitation variability. These coefficients were also present in the top 12.5% (8/64) of best-performing pregnancy models (Figure S3). Posterior distributions for the best-performing models are shown in Figure S4. Posterior estimates for climate lag weights were largely uninformative on the importance of specific climate lags (posterior distribution medians relatively uniform over prior distribution range of 0–168 days; Figure S5).

The model of male population size with the highest predictive accuracy, as indicated by expected log predictive densities, had a random intercept for month; this model outcompeted both simpler and more complex models (Table S3; Eq. 8).

Posterior predictive checks indicated good concordance between predicted and observed values for growth (Figure S6), survival (Figure S7), pregnancy (Figure S8), and male population size (Figure S9).

The posterior distributions of parameters for each demographic process model are shown in Figure S4. Posterior predictions of the contribution of each estimated parameter to the demographic process response is shown in Figure S10. Posterior predictions of the number of pregnancies are shown in Figure S11. Posterior predictions of the proportion of the modelled population in each epidemiological compartment are shown in Figure S12. Posterior predictions of prevalence compared to seroprevalence are shown in Figure S13.

When assessing the ability of the median predicted seroprevalence (see main text) to capture 2017 hold-out data using a Bayesian GLM, we found moderately better predictive ability compared to an intercept-only null model (expected log predictive density difference 13.4 (expected log predictive density difference standard error 11.3)).

The median Lassa fever incubation period was estimated as $\log \mu = 7.50$ (95% CrI $(6.62, 8.55)$) days, with a logarithm of scale parameter $\sigma = 0.30$ $(0.22, 0.42)$. This gives a central 95% prediction interval for the incubation period of $(4.04, 14.08)$ days.

Figure S14 shows a comparison of the seasonal pattern of Lassa fever outbreaks to the seasonal patterns of both *M. natalensis* climatic/demographic drivers and the model-predicted number of infected rodents (Figure S14).

**Table S1. Model parameters and functions.**

| Parameter/function | Definition | Value/prior | Ref. |
|---|---|---|---|
| $\alpha_\mu$ | Intercept for mean of body weight change model | $\mathrm{N}(0, 1)$ | Eqs. 3, 4 |
| $\alpha_\sigma$ | Intercept for variance of body weight change model | $\mathrm{N}(0, 1)$ | Eqs. 3, 4 |
| $\alpha_S$ | Intercept for survival model | $\mathrm{N}(0, 2)$ | Eq. 3 |
| $\alpha_P$ | Intercept for pregnancy model | $\mathrm{N}(0, 3)$ | Eq. 3 |
| $\beta_\mu$ | Coefficient for mean of body weight change model | $\mathrm{N}(0, 1)$ | Eqs. 3, 4 |
| $\beta_\sigma$ | Coefficient for variance of body weight change | $\mathrm{N}(0, 1)$ | Eqs. 3, 4 |
| $\boldsymbol{\beta}_S$ | Vector of coefficients for survival | $\mathrm{N}(0, 1)$ | Eq. 3 |
| $\boldsymbol{\beta}_P$ | Vector of coefficients for pregnancy | $\mathrm{N}(0, 2)$ | Eq. 3 |
| $\boldsymbol{\omega}$ | Climate lag weights for body weight, survival, and pregnancy models | $\mathrm{Dir}(\alpha = 1)$ | Eq. 3, Section B.1 |
| $p_B(y\|w, t)$ | Probability distribution of body weight change | Estimated | Eqs. 3, 5 |
| $p_R(w, t)$ | Probability of recapture | Estimated | Eqs. 3, |
| $p_S(w, t)$ | Probability of survival | Estimated | Eqs. 3, , 6, 7 |
| $p_P(w, t)$ | Probability of pregnancy | Estimated | Eq. 3 |
| $\kappa$ | Probability that an individual captured at time $t$ is still alive at time $t + 1$ despite never being recaptured | Estimated | Eqs. 6, 7 |
| $l$ | No. female offspring per pregnancy | 5 | (3) |
| $v(w)$ | Juvenile weight distribution | $\mathrm{N}(13, 0.3)$ | (11) |
| $\beta$ | Horizontal rate of transmission | $\mathrm{U}(0.00001, 0.0015)$ | Section G |
| $\varphi$ | Probability of vertical transmission | $\mathrm{Beta}(20, 2)$ | (15, 16), Section G |
| $\gamma$ | Recovery rate from horizontal infection | $28^{-1}$ days | (13) |
| $\eta$ | Seroreversion rate of maternal antibodies | $28^{-1}$ days | (14) |

**Table S2. Data collated on nosocomial transmission of Lassa virus, used to estimate an incubation period distribution.**

| Location | Exposure period | Symptom onset period | Incubation period (days) | Fatal (date) | Ref |
|---|---|---|---|---|---|
| Jos, Plateau State, Nigeria | 25/01/1970 | 03/02/1970 | 9 | Yes (13/02/1970) | (21) |
| Jos, Plateau State, Nigeria | 25/01/1970–13/02/1970 | 20/02/1970 | 7–26 | No | (21) |
| Zorzor, Lofa County, Liberia | 02/03/1972–18/03/1972 | 22/03/1972 | 4–20 | No | (22) |
| Zorzor, Lofa County, Liberia | 03/03/1972–19/03/1972 | 22/03/1972 | 3–19 | No | (22) |
| Zorzor, Lofa County, Liberia | 15/03/1972–19/03/1972 | 20/03/1972 | 1–5 | Yes (04/04/1972) | (22) |
| Zorzor, Lofa County, Liberia | 02/03/1972–19/03/1972 | 20/03/1972 | 1–18 | No | (22) |
| Zorzor, Lofa County, Liberia | 01/03/1972–13/03/1972 | 13/03/1972 | 3–12 | NA | (22) |
| Zorzor, Lofa County, Liberia | 01/03/1972–08/03/1972 | 17/03/1972 | 9–16 | NA | (22) |
| Imo State, Nigeria | 23/01/1989 | 29/01/1989 | 6 | Yes (15/02/1989) | (23) |
| Imo State, Nigeria | 25/02/1989 | 03/03/1989 | 6 | Yes (15/03/1989) | (23) |
| Imo State, Nigeria | 25/02/1989 | 03/03/1989 | 6 | Yes (15/03/1989) | (23) |
| Imo State, Nigeria | 25/02/1989 | 07/03/1989 | 10 | No | (23) |
| Imo State, Nigeria | 25/02/1989 | 07/03/1989 | 10 | NA | (23) |
| Abakaliki, Ebonyi State, Nigeria | 03/01/2012 | 08/01/2012 | 5 | NA | (24) |
| Abakaliki, Ebonyi State, Nigeria | 03/01/2012 | 08/01/2012–15/01/2012 | 5–12 | NA | (24) |
| Abakaliki, Ebonyi State, Nigeria | 03/01/2021 | 08/01/2012–15/01/2012 | 5–12 | NA | (24) |
| Abakaliki, Ebonyi State, Nigeria | 03/01/2021 | 08/01/2012–15/01/2012 | 5–12 | NA | (24) |
| Abakaliki, Ebonyi State, Nigeria | 03/01/2021 | 08/01/2012–15/01/2012 | 5–12 | NA | (24) |
| Tanguiétta, Atakora Department, Benin | 07/10/2014–14/10/2014 | 21/10/2014 | 7–14 | Yes (NA) | (25) |
| Tanguiétta, Atakora Department, Benin | 07/10/2014–14/10/2014 | 23/10/2014 | 9–16 | Yes (NA) | (25) |
| Tchaourou, Borgou Department, Benin | 03/01/2016 | 08/01/2016 | 5 | No | (25) |
| Tchaourou, Borgou Department, Benin | 03/01/2016 | 12/01/2016 | 9 | No | (25) |
| Tchaourou, Borgou Department, Benin | 03/01/2016 | 13/01/2016 | 10 | Yes (NA) | (25) |
| Tchaourou, Borgou Department, Benin | 03/01/2016 | 13/01/2016 | 10 | No | (25) |
| Nigeria | 28/12/2017 | 03/01/2018 | 6 | Yes (14/01/2018) | (26) |
| Nigeria | 28/12/2017 | 09/01/2018 | 12 | Yes (14/01/2018) | (26) |
| Nigeria | 28/12/2017 | 03/01/2018 | 6 | Yes (04/01/2018) | (26) |
| Nigeria | 28/12/2017 | 05/01/2018 | 8 | No | (26) |
| Sierra Leone | 04/11/2019 | 11/11/2019 | 7 | Yes (NA) | (27) |
| Sierra Leone | 04/11/2019 | 11/11/2019 | 7 | No | (27) |

**Table S3. Predictive accuracy of four competing Bayesian models of male population size. Models are listed in order of performance. Metrics are in relation to the top performing model, with negative ELPD difference indicating poorer performance and SE differences quantifying uncertainty in ELPD. See Sections B.5 and D for a description of models. ELPD, expected log predictive density. SE, standard error.**

| | ELPD difference | SE difference |
|---|---|---|
| Model 2 | 0.0 | 0.0 |
| Model 4 | 0.0 | 0.2 |
| Model 1 | -49.9 | 10.2 |
| Model 3 | -51.3 | 10.4 |

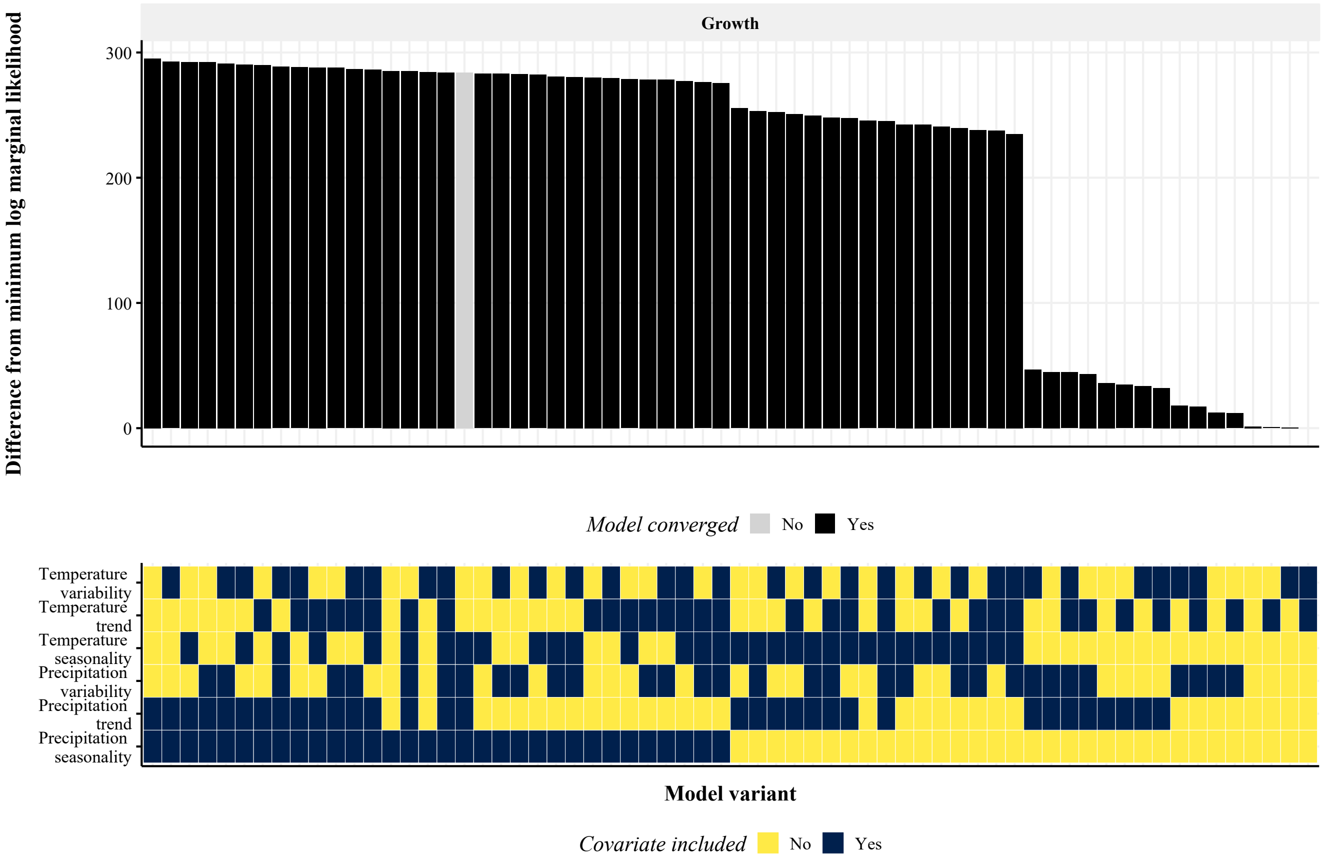


**Fig. S1.** Body weight change (growth) model performance for 64 model variants comprising all combinations of 6 climate predictors. The model estimates body weight at time $t+1$ given demographic and climate covariates at time $t$. Models were fit to three decades of rodent trapping data (3) and climate data (1, 2) using a No-U-Turn Sampler (8) Hamiltonian Monte Carlo algorithm in Stan (7), with prior distributions as specified in Table S1. Approximate marginal log-likelihoods were calculated via bridge sampling (10). The bottom panel shows the covariates included in each model, while the top panel shows the corresponding performance of that model relative to the worst-performing model. Model variants are ordered from best (left) to worst (right) performing. Grey bars indicate models that did not converge ($\hat{R} \geq 1.05$ for $\geq 1$ parameter(s)).

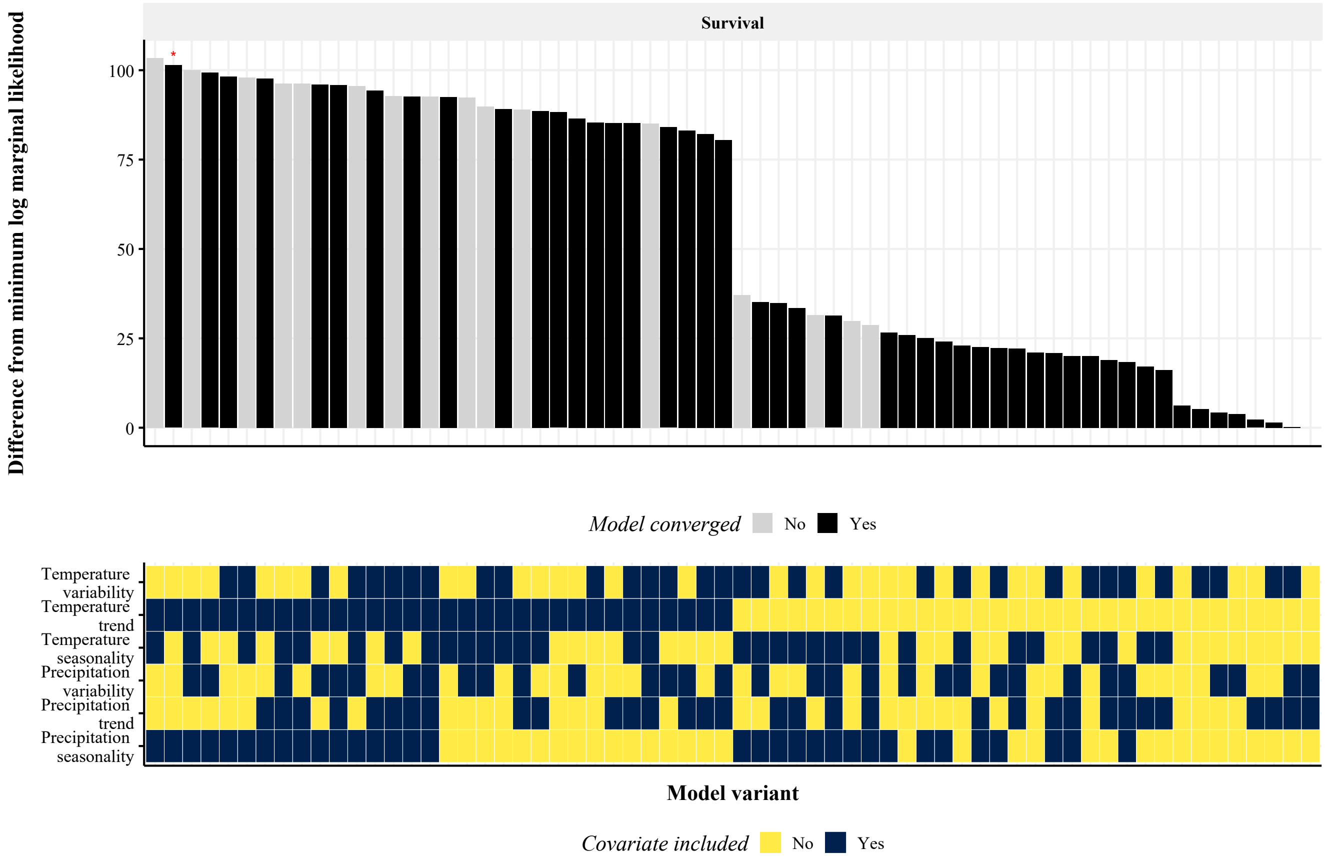


**Fig. S2.** Survival model performance for 64 model variants comprising all combinations of 6 climate predictors. The model estimates recapture at time $t + 1$ given demographic and climate covariates at time $t$. These estimates are subsequently adjusted to obtain probabilities of survival (Section F.4). Models were fit to three decades of rodent trapping data (3) and climate data (1, 2) using a No-U-Turn Sampler (8) Hamiltonian Monte Carlo algorithm in Stan (7), with prior distributions as specified in Table S1. Approximate marginal log-likelihoods were calculated via bridge sampling (10). The bottom panel shows the covariates included in each model, while the top panel shows the corresponding performance of that model relative to the worst-performing model. Model variants are ordered from best (left) to worst (right) performing. Grey bars indicate models that did not converge ($\hat{R} \geq 1.05$ for $\geq 1$ parameter(s)). *Bayes factor $\geq 5$ and $< 10$ compared to the best-performing model.

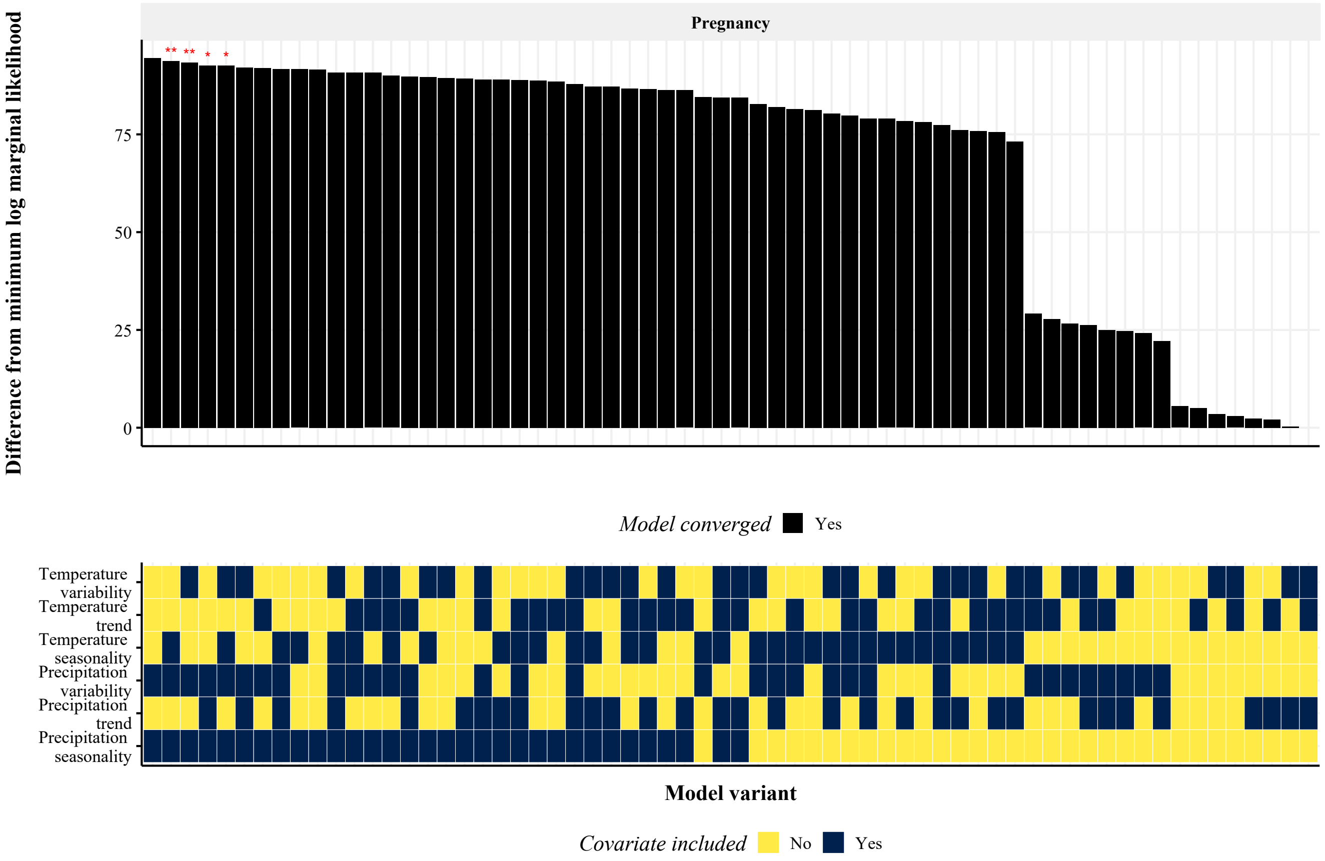


**Fig. S3.** Pregnancy model performance for 64 model variants comprising all combinations of 6 climate predictors. The model estimates pregnancy at time $t + 1$ given demographic and climate covariates at time $t$. Models were fit to three decades of rodent trapping data (3) and climate data (1, 2) using a No-U-Turn Sampler (8) Hamiltonian Monte Carlo algorithm in Stan (7), with prior distributions as specified in Table S1. Approximate marginal log-likelihoods were calculated via bridge sampling (10). The bottom panel shows the covariates included in each model, while the top panel shows the corresponding performance of that model relative to the worst-performing model. Model variants are ordered from best (left) to worst (right) performing. Grey bars indicate models that did not converge ($\hat{R} \geq 1.05$ for $\geq 1$ parameter(s)). **Bayes factor $< 5$ compared to the best-performing model; *Bayes factor $\geq 5$ and $< 10$ compared to the best-performing model.

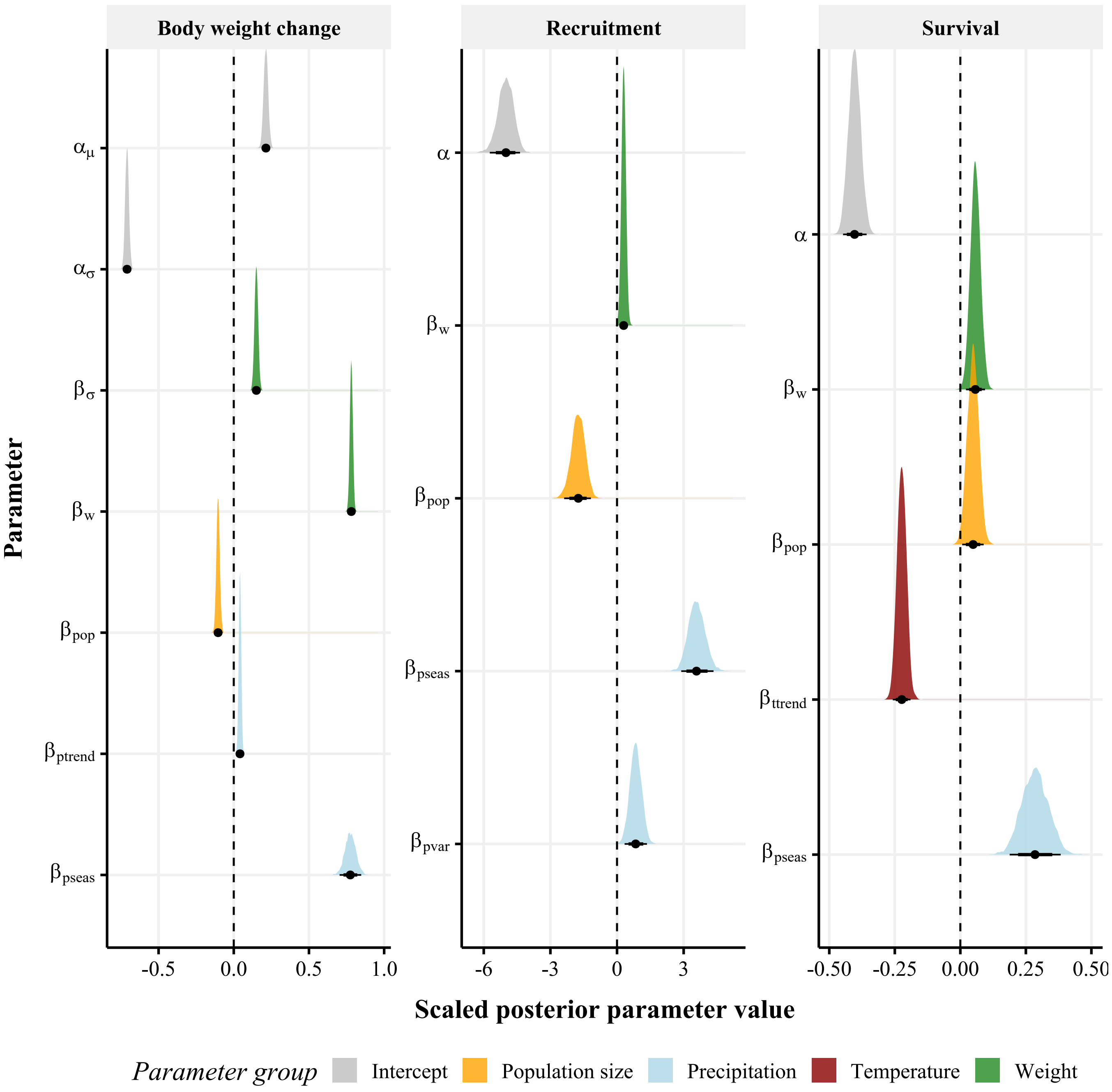


**Fig. S4.** Estimated posterior distributions of unit-scaled coefficients of demographic process models. The density of each posterior distribution is shown alongside a posterior median (point), 80% credible interval (thick horizontal line), and 95% credible interval (thin horizontal line). The vertical dashed line represents no effect. An approximate Bayesian approach was used to fit the model to rodent trapping data (3) and climate data (1, 2). The $\alpha$ parameters represent intercepts for: the Bernoulli processes of recruitment and survival ($\alpha$), the mean of the body weight distribution ($\alpha_\mu$), and the standard deviation of the body weight distribution ($\alpha_\sigma$). The $\beta$ parameters represent coefficients for: the standard deviation of the body weight distribution ($\beta_\sigma$), body weight ($\beta_w$), population size ($\beta_N$), precipitation trend ($\beta_{p_{\mathrm{trend}}}$), precipitation seasonality ($\beta_{p_{\mathrm{seas}}}$), precipitation variability ($\beta_{p_{\mathrm{var}}}$), and temperature trend ($\beta_{t_{\mathrm{trend}}}$).

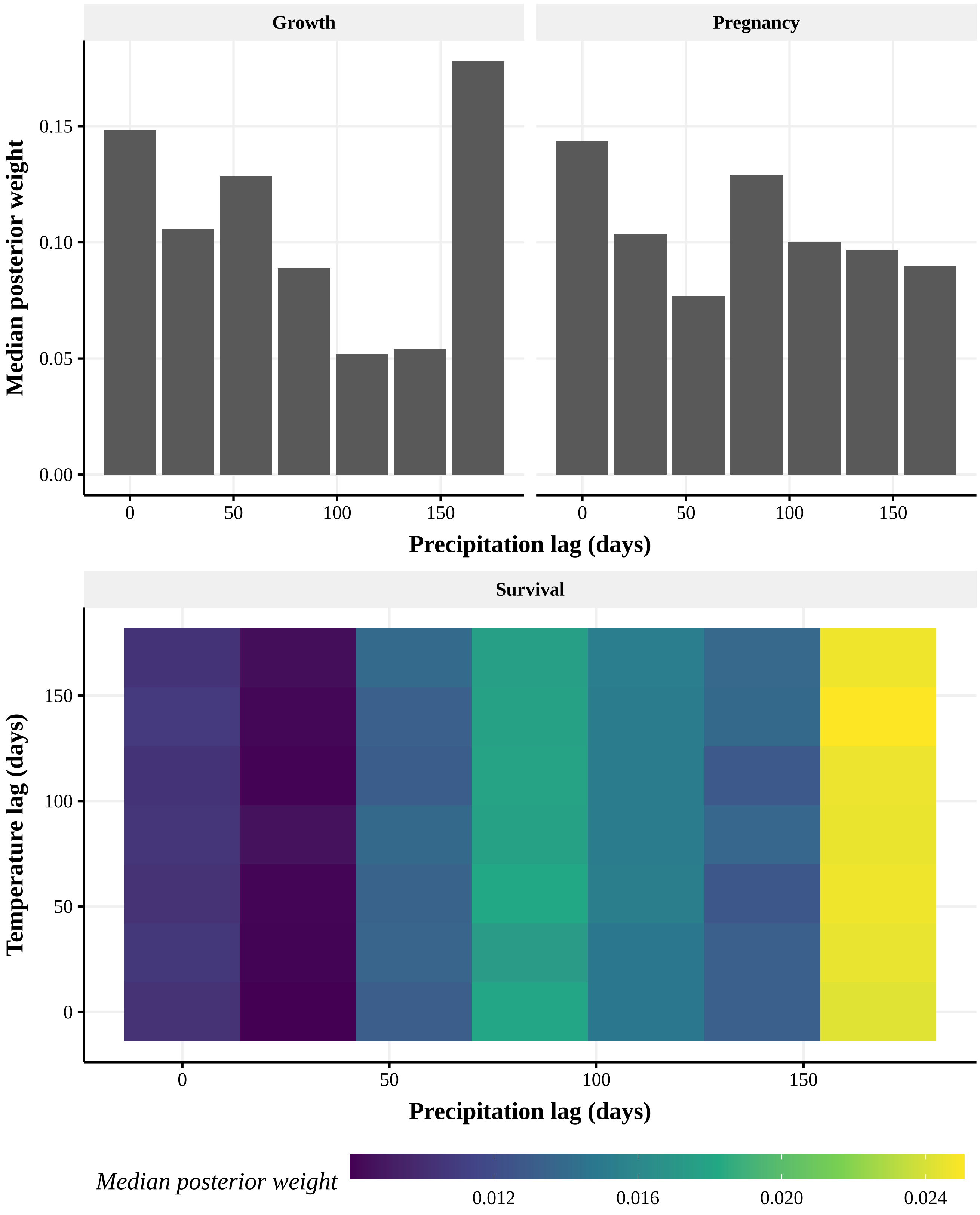


**Fig. S5.** Median posterior estimates of climate lag weights ($N = 4000$ posterior samples) for the best-performing models of growth, pregnancy, and survival. While growth and pregnancy models only included precipitation predictors, survival also included temperature predictors and hence for this model lag weight estimates are shown by precipitation and temperature lag. Models were fit to three decades of rodent trapping data (3) and climate data (1, 2) using a No-U-Turn Sampler (8) Hamiltonian Monte Carlo algorithm in Stan (7), with prior distributions as specified in Table S1.

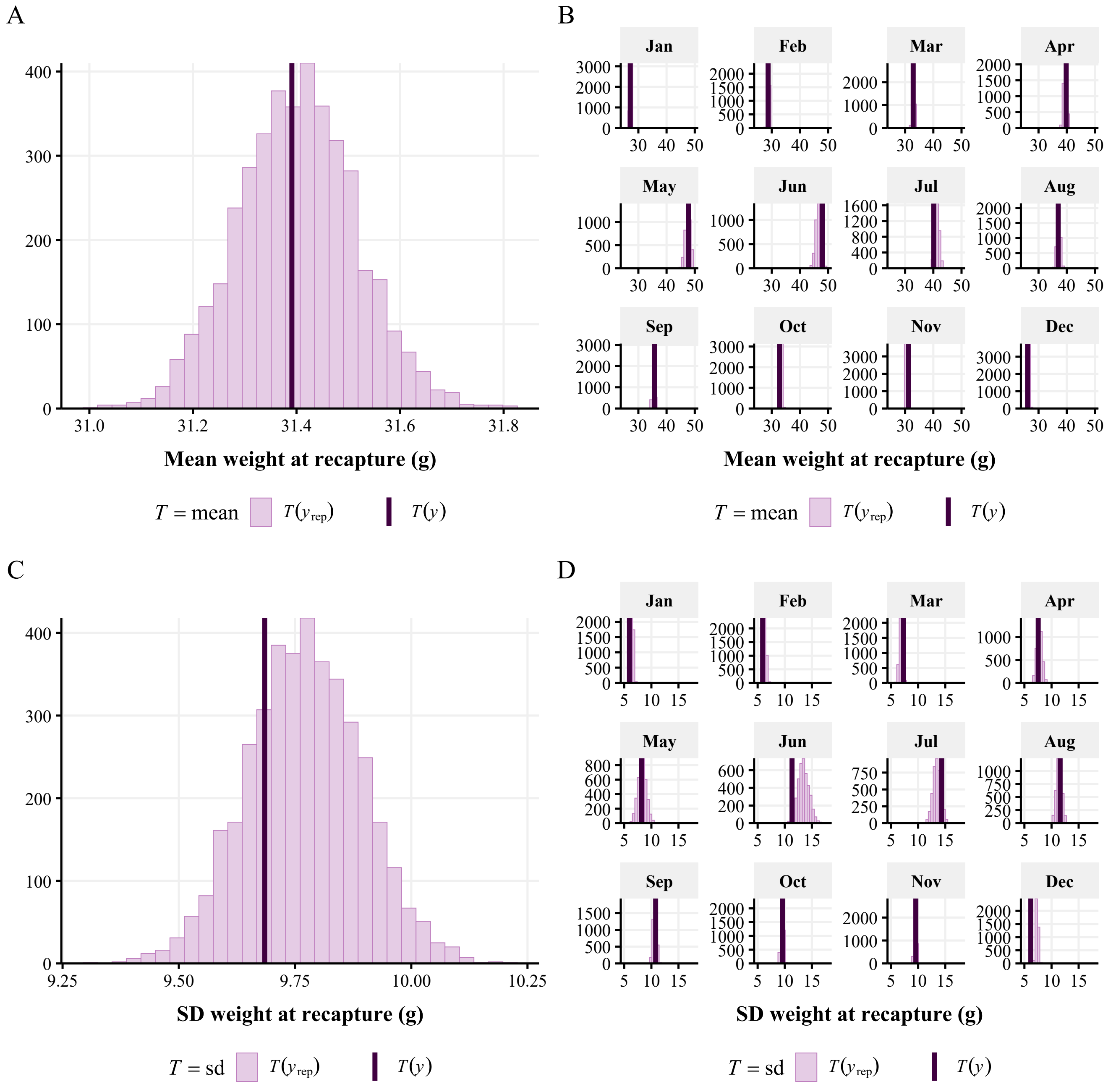


**Fig. S6.** Posterior predictive checks (PPCs) of the best-performing body weight change model. PPCs were performed by simulating $4000$ posterior samples from the posterior predictive distribution of the fitted model to obtain predicted replications $y_{\text{rep}}$ (pink histogram) of the observed data $y$ (dark vertical line). Panels **A** and **B** show for the whole population and by month, respectively, the observed versus predicted values of the mean weight at time $t+1$. Panels **C** and **D** show for the whole population and by month, respectively, the observed versus predicted values of the standard deviation of weight at time $t+1$.

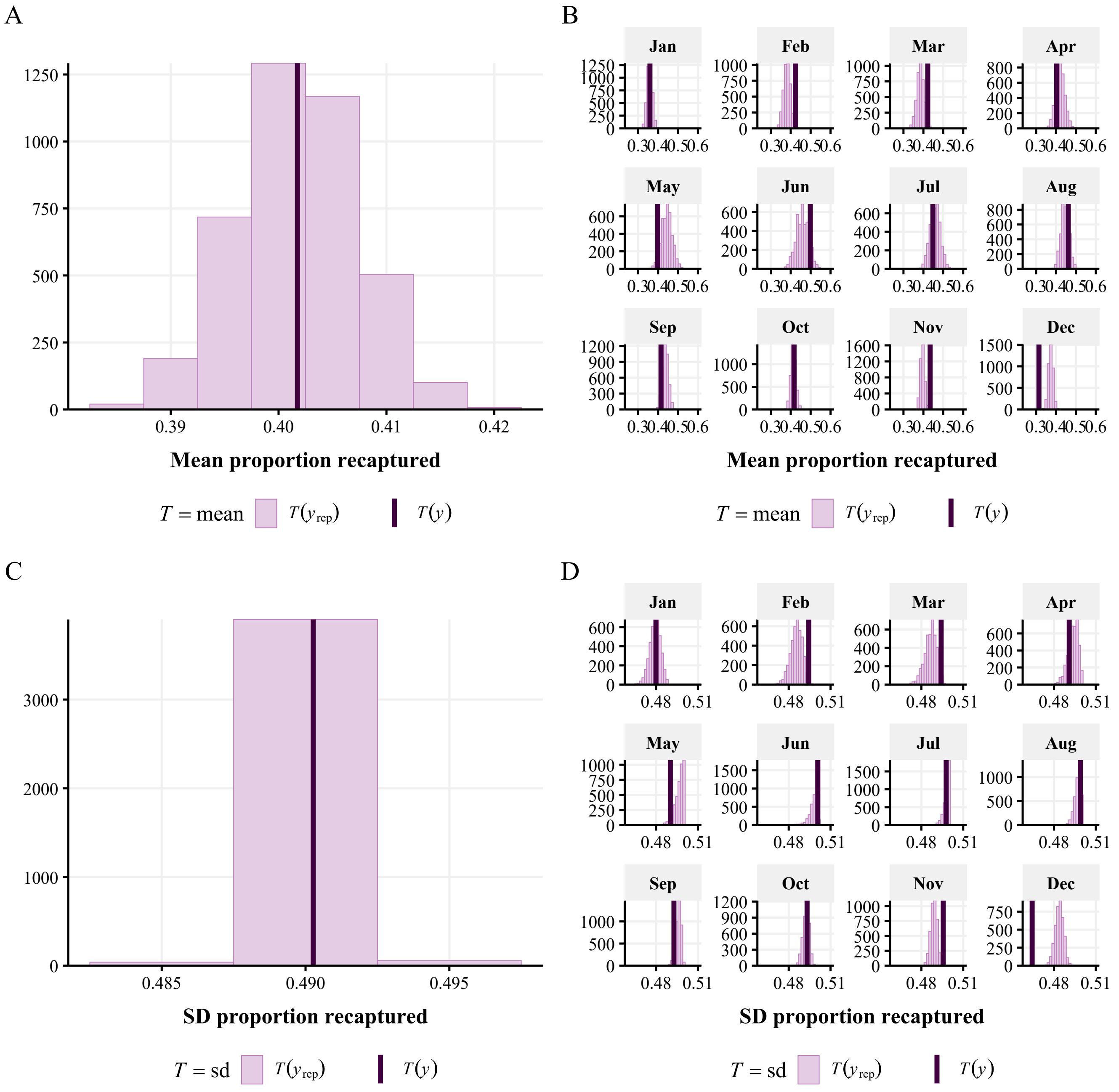


**Fig. S7.** Posterior predictive checks (PPCs) of the best-performing survival model. PPCs were performed by simulating $4000$ posterior samples from the posterior predictive distribution of the fitted model to obtain predicted replications $y_{\text{rep}}$ (pink histogram) of the observed data $y$ (dark vertical line). Panels **A** and **B** show for the whole population and by month, respectively, the observed versus predicted values of the mean proportion of individuals recaptured at time $t+1$. Panels **C** and **D** show for the whole population and by month, respectively, the observed versus predicted values of the standard deviation of the proportion of individuals recaptured at time $t+1$.

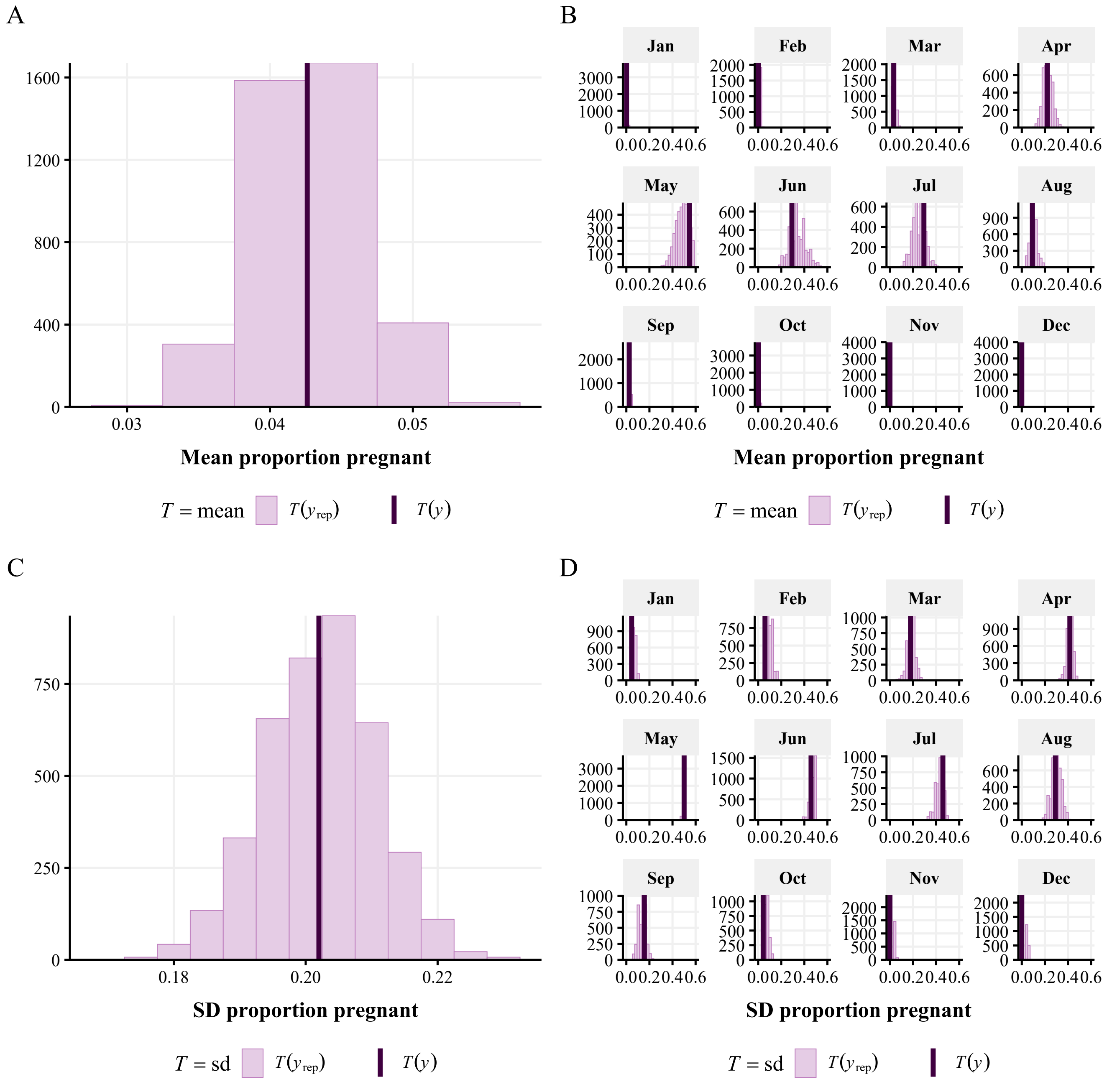


**Fig. S8.** Posterior predictive checks (PPCs) of the best-performing pregnancy model. PPCs were performed by simulating $4000$ posterior samples from the posterior predictive distribution of the fitted model to obtain predicted replications $y_{\text{rep}}$ (pink histogram) of the observed data $y$ (dark vertical line). Panels **A** and **B** show for the whole population and by month, respectively, the observed versus predicted values of the mean proportion of individuals pregnant at time $t + 1$. Panels **C** and **D** show for the whole population and by month, respectively, the observed versus predicted values of the standard deviation of the proportion of individuals pregnant at time $t + 1$.

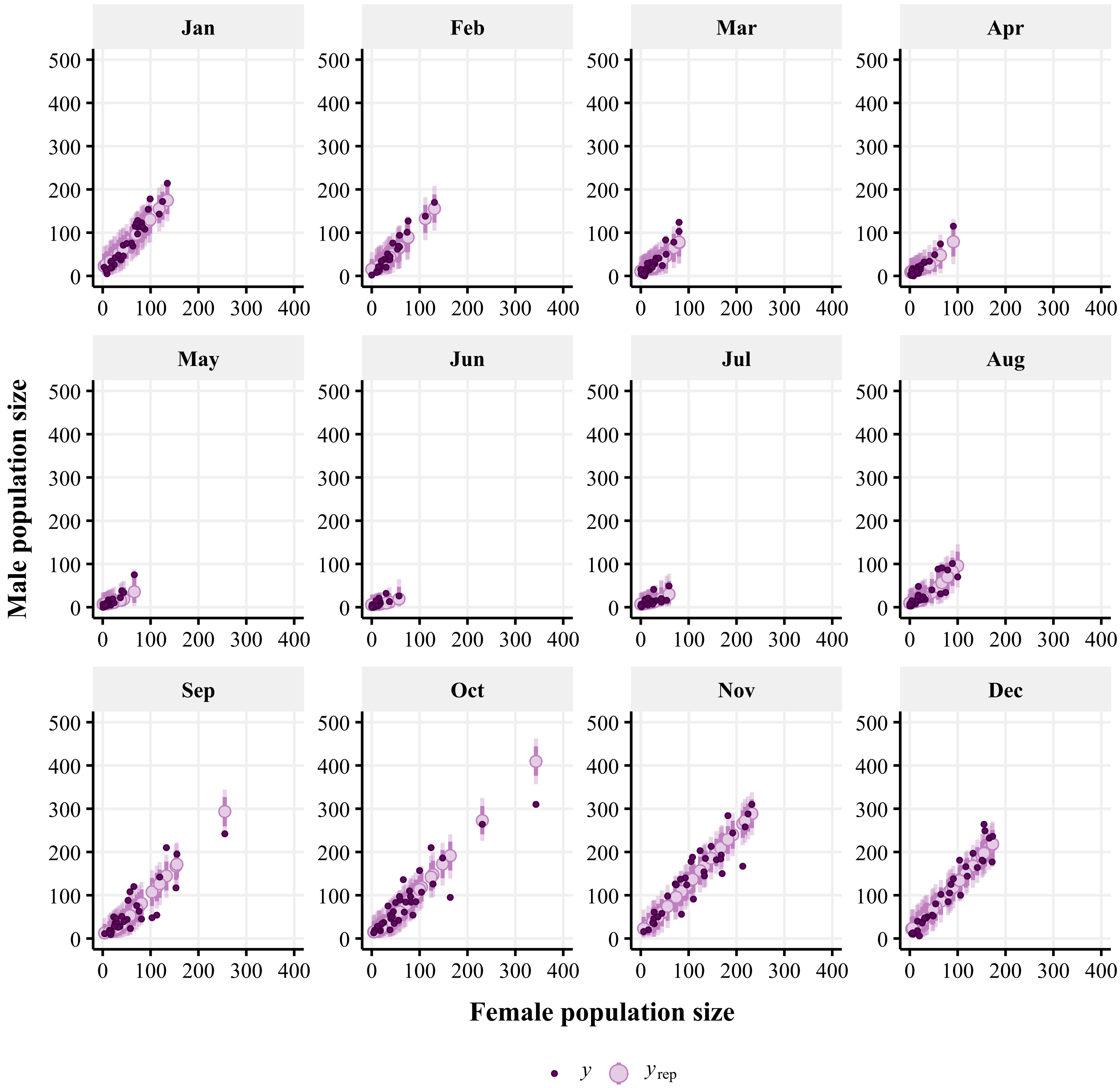


**Fig. S9.** Posterior predictive checks (PPCs) of the best-performing male population size model, which includes a coefficient for female population size and month as a random intercept. PPCs were performed by simulating $4000$ posterior samples from the posterior predictive distribution of the fitted model to obtain predictions $y_{\mathrm{rep}}$ (pink points representing medians and inner and outer error bars 80% and 95% credible intervals, respectively) of the observed data $y$ (dark points).

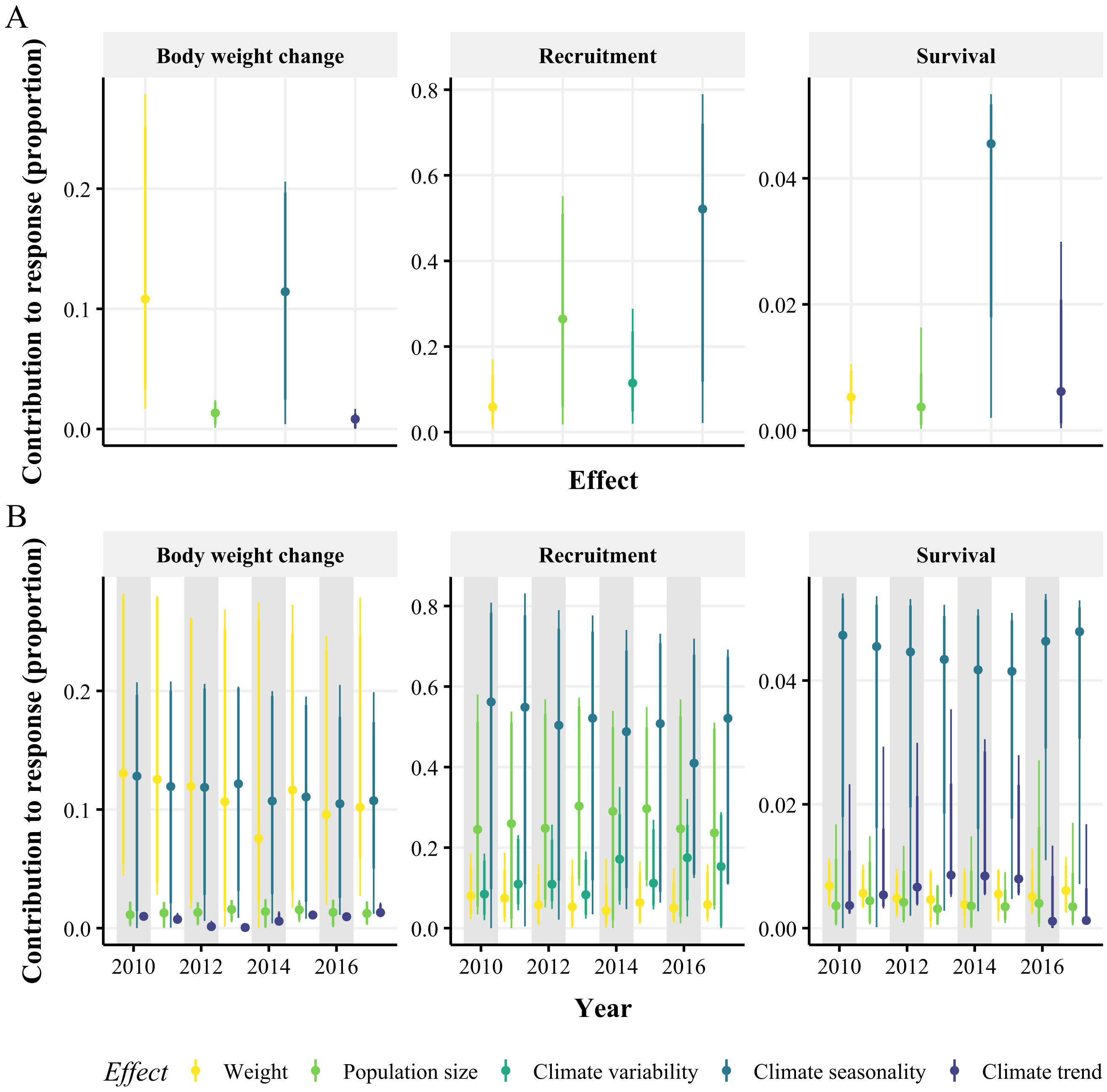


**Fig. S10.** Posterior predictions ($N = 1000$ simulations) of the contribution of each covariate to the modelled demographic process (shown as a proportion of the total response over time (**A**) and the year-specific proportion of the response (**B**)). The median prediction (point) is shown alongside the 80% credible interval (thick vertical line), and 95% credible interval (thin vertical line). An approximate Bayesian approach was used to fit the model to rodent trapping data (3), climate data (1, 2), and arenavirus serological data (14). Posterior predictive samples were obtained through Bayesian melding (Section H) (17, 18).

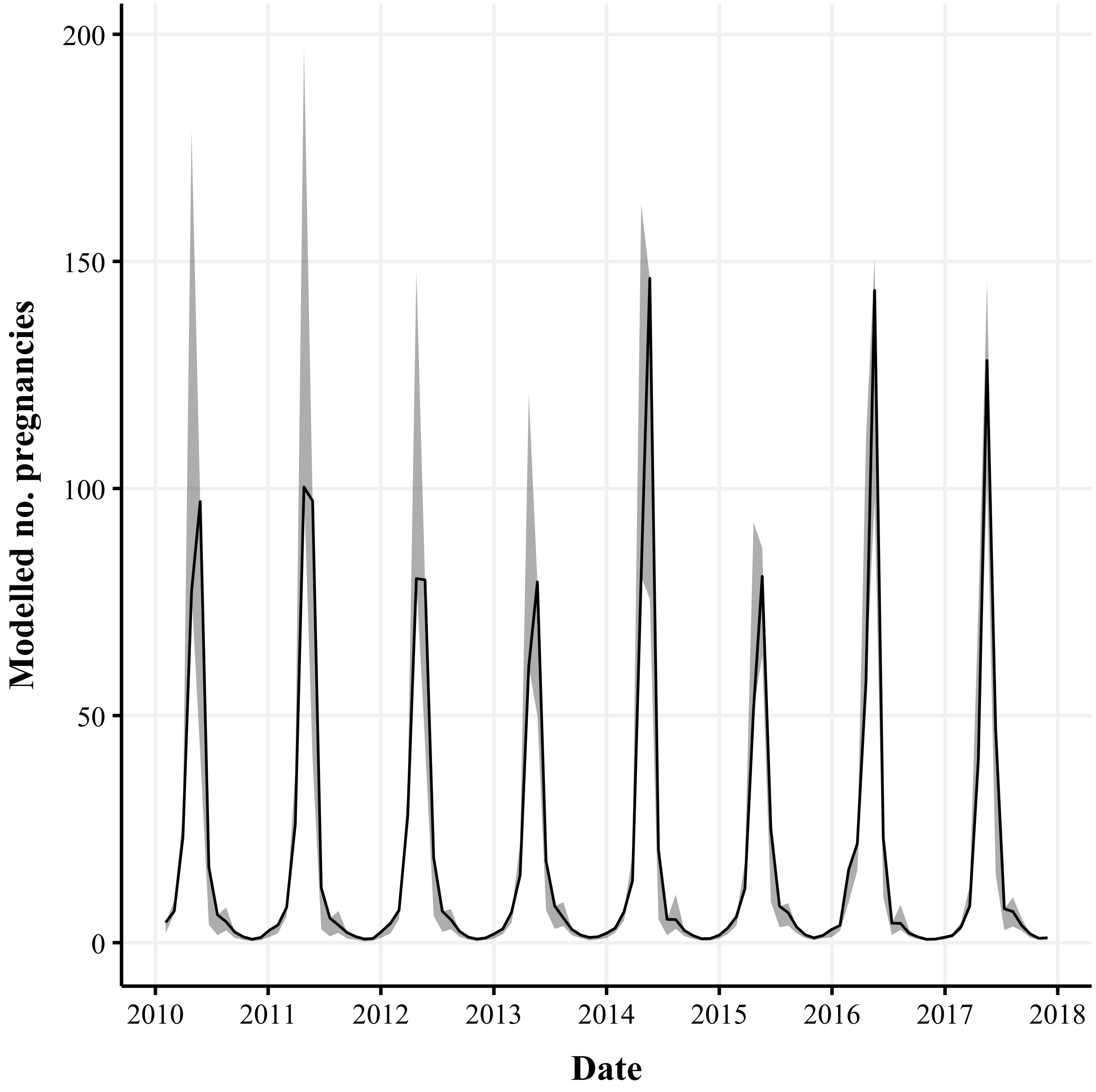


**Fig. S11.** Posterior predictions ($N = 1000$ simulations) of the predicted number of pregnant rodents. The line represents the median while shaded areas represent the 95% credible interval. An approximate Bayesian approach was used to fit the model to rodent trapping data (3), climate data (1, 2), and arenavirus serological data (14). Posterior predictive samples were obtained through Bayesian melding (Section H) (17, 18).

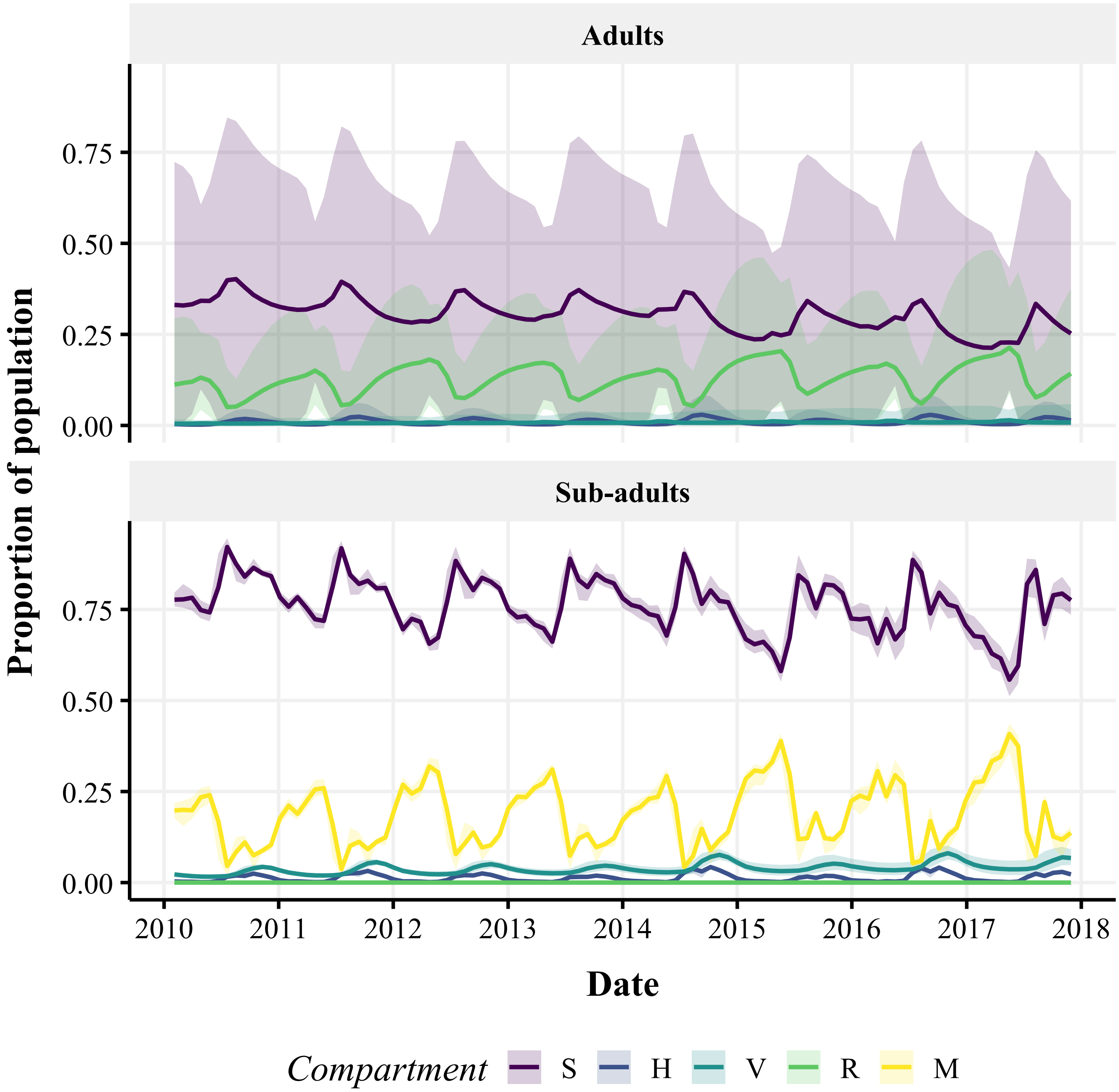


**Fig. S12.** Posterior predictions ($N = 1000$ simulations) of the proportion of the modelled population in each epidemiological compartment at a given time: susceptible ($S$), horizontally infected ($H$), vertically infected ($V$), recovered ($R$), and maternal antibody positive ($M$). The time-specific proportion of the population in each compartment was calculated for each model time-step by demographic group (sub-adult vs. adult) independently for each posterior simulation. Lines represent the medians while shaded areas represent 95% credible intervals. An approximate Bayesian approach was used to fit the model to rodent trapping data (3), climate data (1, 2), and arenavirus serological data (14). Posterior predictive samples were obtained through Bayesian melding (Section H) (17, 18).

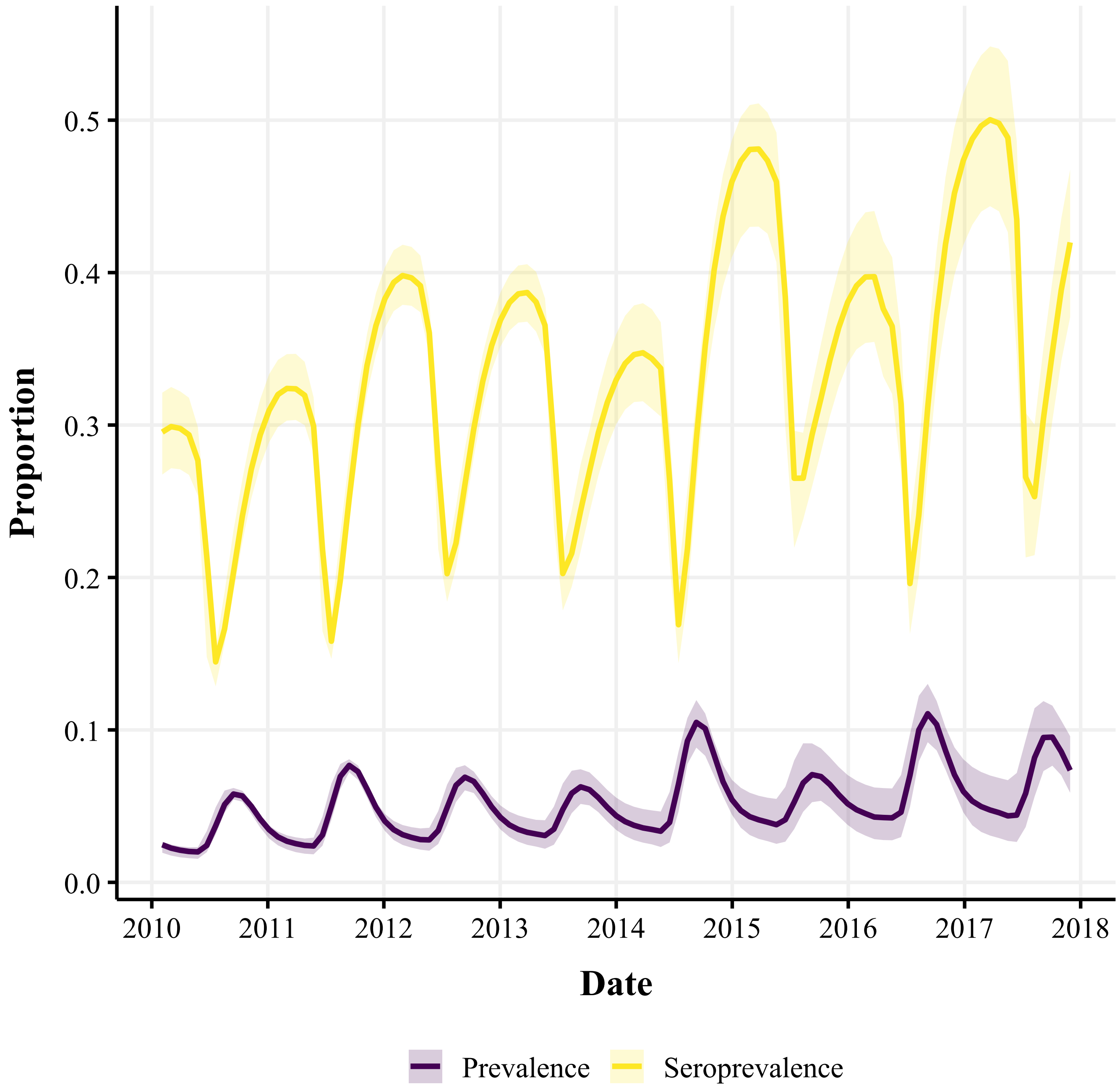


**Fig. S13.** Posterior predictions ($N = 1000$ simulations) of the relationship between prevalence and seroprevalence. Lines represent the medians while shaded areas represent 95% credible intervals. The time-specific prevalence was calculated as $[H(t) + V(t)]/N(t)$. The time-specific seroprevalence was calculated as $[N(t) - S(t)]/N(t)$ (Section G). An approximate Bayesian approach was used to fit the model to rodent trapping data (3), climate data (1, 2), and arenavirus serological data (14). Posterior predictive samples were obtained through Bayesian melding (Section H) (17, 18).

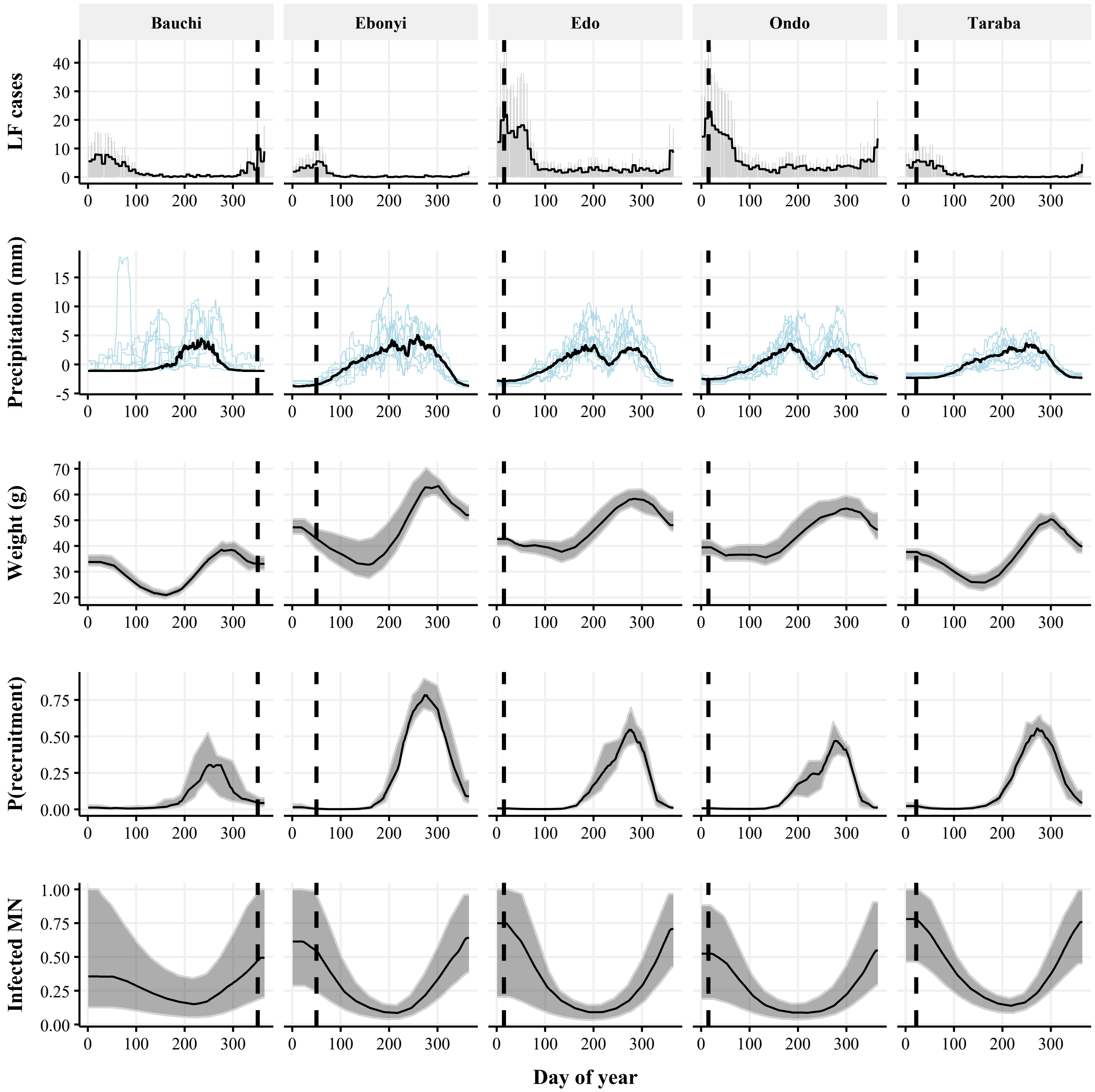


**Fig. S14.** Comparison for five Nigerian states of the seasonal pattern of Lassa fever (LF) outbreaks (top row) to those of the climatic and demographic drivers of *Mastomys natalensis* (MN) population dynamics (middle three rows), and of the model-predicted number of infected MN (bottom row). The top row of panels shows weekly laboratory-confirmed LF cases extracted 2018–2025 (19), both as a seasonal mean (black line) and as the raw data (grey bars). The black vertical dashed line indicates the average timing of the highest case numbers for that state. The second row of panels shows detrended precipitation data (1, 2) as the seasonal (black line) and variable (blue lines) components from a decomposed time series. The bottom three rows of panels (MN weight, probability of recruitment, and the relative number of infected MN) show the median (black line) and 95% credible interval (grey shaded area) from $N = 1000$ posterior predictive simulations over 2018–2025. For each simulation, weight and P(recruitment) were calculated as time-specific means across the population. The number of infected MN was calculated as $H(t) + V(t)$ and made relative to the maximum value in each simulation before calculating summarised values across all simulations. An approximate Bayesian approach was used to fit the model to rodent trapping data (3), climate data (1, 2), and arenavirus serological data (14). Posterior predictive samples were obtained through Bayesian melding (Section H) (17, 18).
